# Sex-different phenotypic correlations: Due to genes or environment?

**DOI:** 10.64898/2026.07.13.26357694

**Authors:** Amelie Fritz, Liza Darrous, Klaus Bønnelykke, Anders G. Pedersen, Zoltán Kutalik

## Abstract

Differences in physical features and disease prevalence between men and women are examples of sexual dimorphisms. However, sex differences can manifest not only in trait means but also in how strongly risk factors are linked to diseases (e. g. BMI to cardiovascular disease), a question heavily under-researched. To fill this gap, we set out to identify sex differences in phenotype correlations (rP) and decompose them into genetic (rG) and environmental (rE) contributions.

Our analysis revealed 250 trait pairs with significant sex-different phenotypic correlations in the UK Biobank. Overall, we observed a predominance of environmental contributions to sex-different effects: 182 trait pairs (73%) exhibited exclusively sex-different rE, while 68 (27%) showed sex differences in both rE and rG, and no trait pair was affected solely by sex-specific rG. For example, we detected sex-different environmental correlation between C-reactive protein and BMI (rE(men) = 0.07 *vs* rE(women) = 0.25), but no sex-difference in genetic correlation. On the contrary, glycated haemoglobin and LDL cholesterol showed genetic correlation only in women (rG(women) = 0.17; 95% CI = [0.1, 0.23]), but environmental correlation only in men (rE(men) = -0.18; 95% CI = [-0.19, -0.16]). Some of the observed sex differences – including those involving testosterone, SHBG, urate, waist-hip ratio, and triglycerides – may reflect underlying sex-specific genetic architectures, as evidenced by low between-sex genetic correlations.

In conclusion, environmental factors are the predominant contributors to sex differences in phenotypic correlations between complex traits, with modest detectable contributions from sex-specific genetic architectures. Recognising these patterns can inform the development of more effective, sex-informed interventions.

## Introduction

Sexual dimorphism is evident and measurable in numerous physical and clinical traits, including disease prevalence, onset, and severity, as well as anatomical, physiological, psychiatric, behavioural traits, and mental disorders, and disorders of the immune system (1,2). For example, females are more prone to be affected by autoimmune diseases (3,4), anxiety (5), and depression (6); while males are found to be at higher risk for heart disease and Type 2 diabetes (T2D) compared to women up to menopausal age, after which the risk for both diseases start to increase (7,8). Childhood asthma is more prevalent amongst boys, while after puberty, more women than men are reported to have asthma.

Sexual dimorphism manifests itself not only in the mean value of a trait, but also in sex-different correlations between traits. Recent studies have shown that lipid markers and depression have a stronger association with cardiovascular risk in men than in women, while eating habits have a stronger influence on cardiovascular risk in women than in men (11).

While there is evidence for sex-specific genetic effects (1,12,13), it often remains challenging to pinpoint the specific mechanisms involved (14). In a review, Patsopoulos *et al.* looked at 215 published studies that claimed to have found sex-specific genetic effects and found that most sex-specific genetic effects were in fact spurious or insufficiently documented, most likely due to very small sample sizes after subgrouping the data set for males and females (15).

Phenotypic correlations between two traits may arise from various factors, including uni- or bi-directional causal effects or shared confounders. If these correlations differ between men and women, such an observation translates to important consequences for disease aetiology and personalised medicine. Sex-specific correlations may reflect interactions with hormonal levels or represent sex-specific environmental exposures (16). Complex traits are influenced by both genetic and environmental factors, which may potentially interact (17). Therefore, phenotypic correlations can also emerge due to genetic and/or environmental reasons. The phenotypic correlation (rP) can thus be decomposed into genetic correlation (rG) and environmental correlation (rE). However, very little is known about which trait pairs exhibit sex differences in rP, and in such cases, whether those differences arise due to genetic or environmental causes.

In this study, we investigated sex differences in phenotypic correlations for 34 traits in the UK Biobank by decomposing them into sex-specific genetic (rG) and environmental (rE) components. To further elucidate sex-specific genetic architecture, we also estimated sex-stratified heritability and cross-sex rG between males and females within individual traits.

## Methods

### Data

We extracted phenotype data from the UK Biobank (18) for 34 different traits measured in unrelated, white individuals of European ancestry, with sample sizes ranging from 51,218 to 337,423 depending on the specific trait (see **Supplementary Table 1**). The participants were between 40 and 69 years old.

We selected traits showing sex-different phenotypic correlations with at least one other trait. The traits can be grouped into the following categories:

**Physical measurements:** standing height (Height), body mass index (BMI), waist-hip ratio (WHR).

**Behavioural traits:** smoking status (smoking: current, never, previous), educational attainment (Edu), alcohol consumption (alcohol).

**Diseases:** asthma, Chronic Obstructive Pulmonary Disease (COPD), Gastroesophageal reflux disease (GERD).

**Lung function measurements:** forced expiratory volume in one second (FEV1), forced vital capacity (FVC).

**Biochemical markers:** Alkaline Phosphatase (ALP), gamma glutamyl transferase (GGT), apoliprotein B (ApoB), cholesterol, low-density lipoprotein cholesterol (LDL), high-density lipoprotein cholesterol (HDL), triglycerides (TG), basophil count, eosinophil count, neutrophil count, cystatin C (CysC), glucose level, glycated haemoglobin (HbA1c), Vitamin D (VitD), C-reactive protein (CRP), haemoglobin (Hb), insulin-like growth factor 1 (IGF1), sex-hormone binding globulin (SHBG), testosterone, oestradiol, urate.

Detailed trait abbreviations, the number of combined, male, and female individuals, and UK Biobank Field IDs are provided in **Supplementary Table 1**.

For each of these 34 traits, we calculated phenotypic correlations (rP), genetic correlations (rG), and environmental correlations (rE) (**Figure 1**).

**Figure 1:**
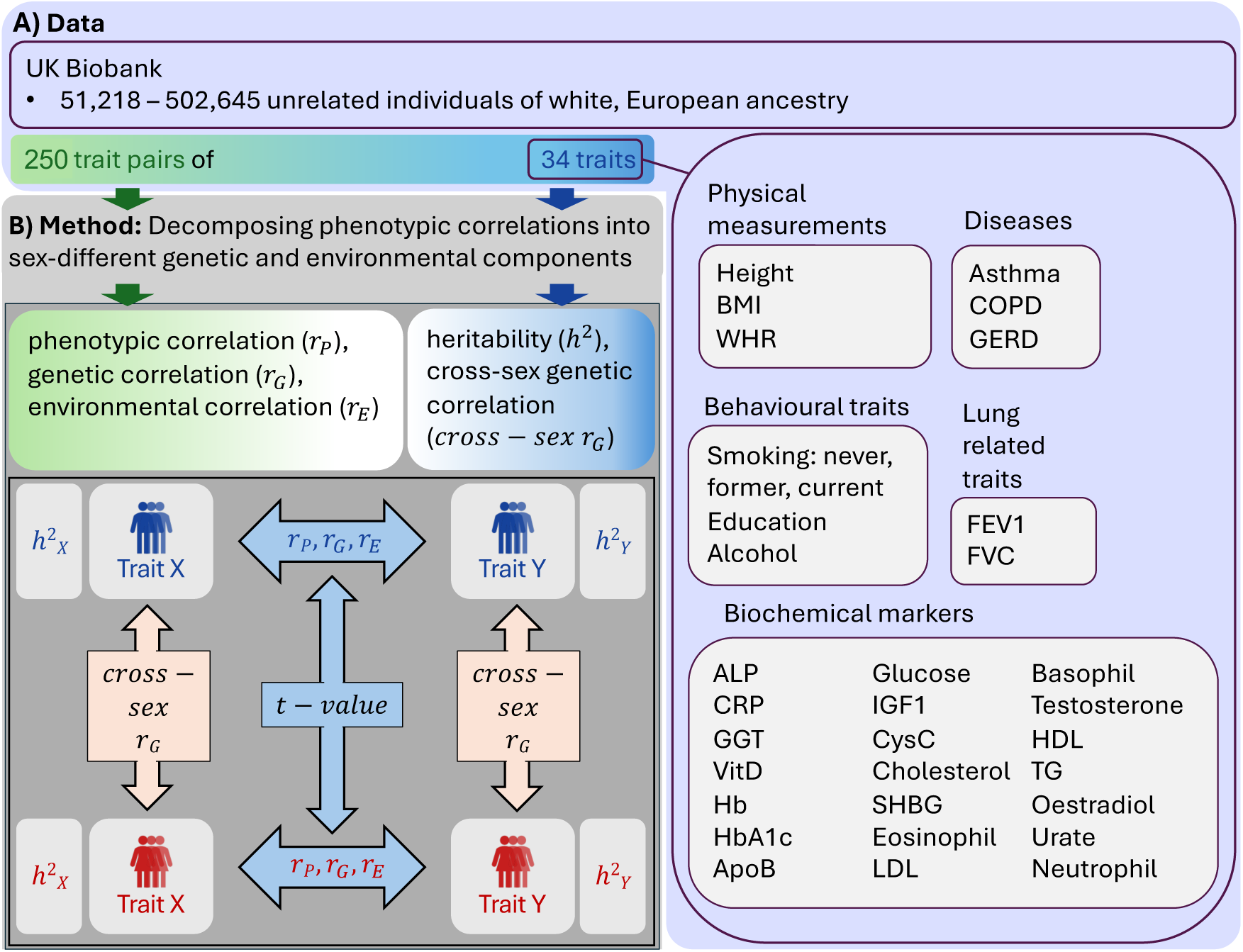
Overview of data (A) and statistical analysis (B). A) We selected 34 traits from the UK Biobank, consisting of 51,218 to 502,645 unrelated individuals of white, European ancestry. B) We decomposed the phenotypic correlations into genetic and environmental contributions. We also calculated the sex-stratified heritability (ℎ^2^), and the cross-sex genetic correlation (*cross* − *sex r_G_*) of the 34 traits. Additionally, to the sex-stratified phenotypic correlation (*r_P_*), we calculated the sex-stratified genetic (*r_G_*) and the sex-stratified environmental correlation (*r_E_*) of the 250 trait pairs. Finally, we determined trait-pairs with sex-different phenotypic, genetic, or environmental correlation. Men are depicted in blue and women in red.

### Correlation analysis

#### Phenotypic correlation

We wanted to investigate the extent to which differences in phenotypic correlations between males and females could be explained by genetic or environmental differences between the sexes.

We selected 34 traits from the UK Biobank and calculated all pairwise phenotypic correlations separately for males and females, using Pearson’s correlation coefficient (19,20):

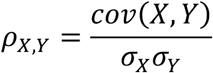

Here, *ρ_X_*_,F_ represents the phenotypic correlation, *cov* the covariance, and *σ_X_* and *σ*_F_ denote the standard deviations of the traits *X* and *Y* respectively.

We assessed whether each pairwise phenotypic, genetic, and environmental correlation significantly differed between males and females by performing a *t*-test comparing the two correlation coefficients (21) (**Figure 1 and Figure 2**).

**Figure 2:**
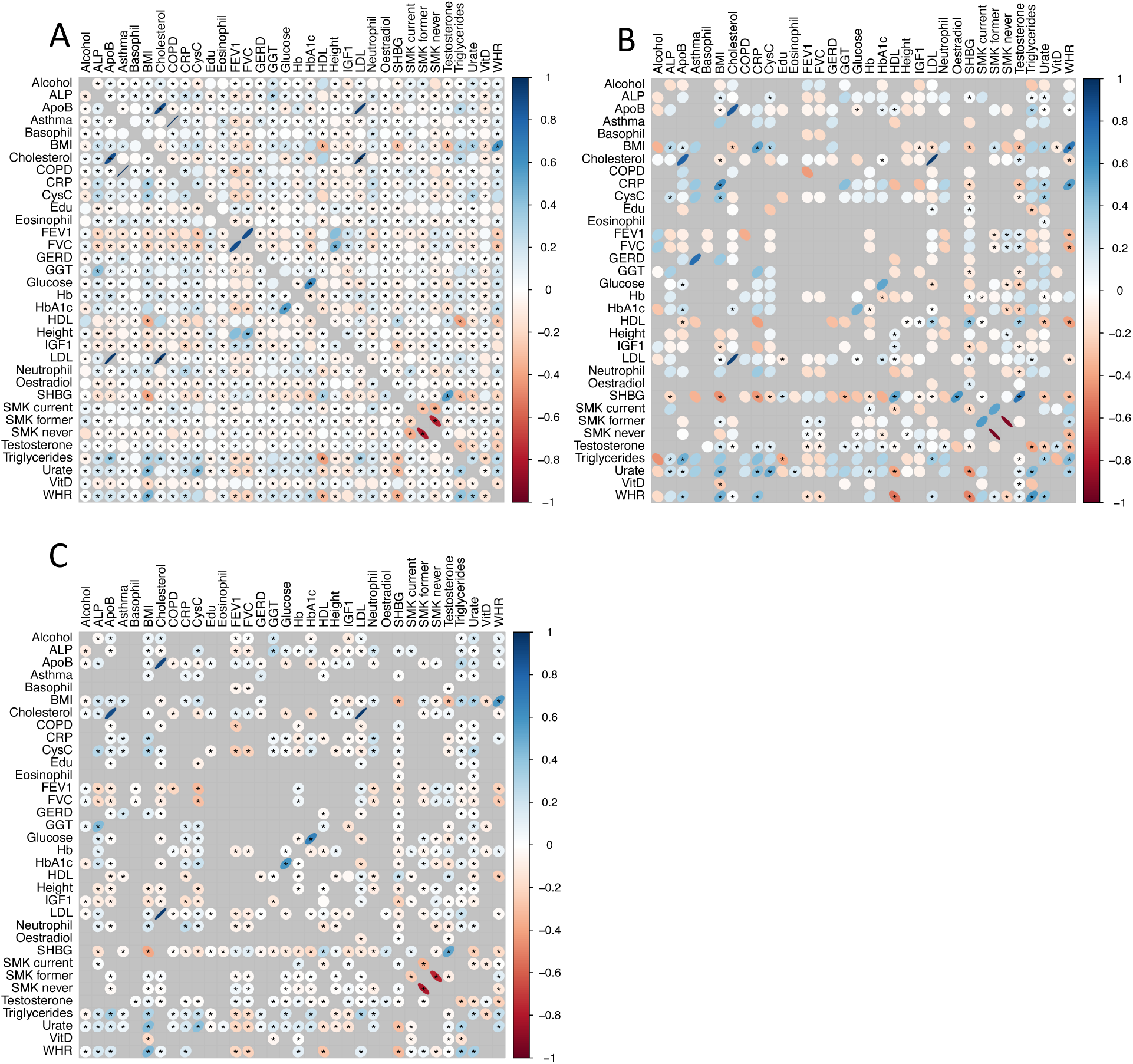
Sex-stratified correlations, upper triangle: women, lower triangle: men. Sex-stratified phenotypic (A), genetic (B), and environmental (C) correlation. Sex-different correlations are marked with an asterisk if FDR < 0.05 (Benjamini– Hochberg corrected). Sex difference was determined by a *t*-test between the respective male and female correlations. For better visibility, values in the heatmaps are signed log-scaled.

#### Genetic correlation

Genetic correlations (*r_G_*) were calculated using LD score regression (22) as implemented in the Genomic Structural Equation Modelling (genomicSEM) R package (23) using R version 4.3.1, 2023-06-16, and version 4.2.2, 2022-10-31. This method estimates genetic correlations based on SNP (single nucleotide polymorphism) heritability derived from Genome-wide Association Studies (GWAS) summary statistics for individual traits, sourced from samples with diverse and unspecified levels of overlap. We used summary statistics provided by the Neale Lab (24) for 33 traits, and computed these ourselves for waist-to-hip ratio (WHR). The Neale lab used the first 10 principal components as covariates (24), while for WHR (unadjusted for BMI), we included the first 40 principal components and genotype batch as covariates.

For the binary traits asthma, COPD, GERD, and smoking (current, never, previous), we transformed the heritability estimations to the liability scale using population prevalence estimates.

#### Environmental Correlation

We calculated the environmental correlation *r_E_* following the approach described by Sodini *et al.* (25). First, we assumed each trait (X, Y) is scaled to have zero mean and unit variance, and that its variance can be decomposed into a genetic component (G) and a residual component representing environmental effects (E):

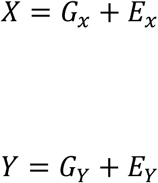

The phenotypic correlation *r_P_* between traits X and Y can then be written as:

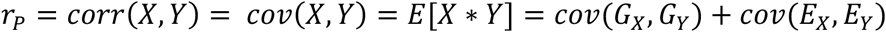

Since both traits are standardised, their covariance equals their correlation (sd=1). This formulation assumes that there is no covariance between genetic and environmental effects across traits: *cov*(*G_X_*, *E*_F_) = 0 and *cov*(*G*_F_, *E_X_*) = 0. If these terms were non-zero, it would imply that part of the genetic influence is still captured within the residual (environmental) component, which contradicts the assumption that E represents only non-genetic variance.

Each covariance term can be expressed as the product of correlations and their respective standard deviations. Thus, the phenotypic correlation can be rewritten as:

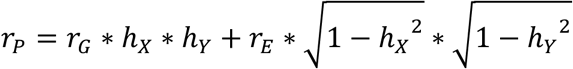

where ℎ*_X_* and ℎ_F_ denote the narrow-sense heritability of the two traits, respectively (26). For the binary traits asthma, COPD, GERD, T2D, and smoking behaviour we used population prevalences (**Supplementary Table 3**) to estimate heritabilities on the liability scales from the observed scales.

Solving this equation for *r_E_* we obtain (25):

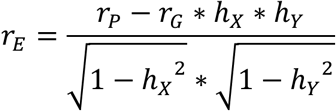

### Establishing sex-differences

To assess whether male- and female-specific estimates (such as heritability, rP, rG, and rE) were significantly different from each other, and to evaluate the difference between genetic and environmental correlations (rG and rE) we used the following test statistic:

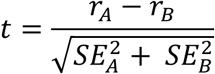

where *r_A_* represents the male-specific estimate (phenotypic, genetic, or environmental correlation), *r_B_* represents the corresponding female-specific estimate, and *SE_A_* and *SE_B_* their respective standard errors. Furthermore, to investigate whether the genetic architecture of a trait differed between men and women, we tested if the cross-sex genetic correlation rG for each trait was significantly different from 1.

## Results

### Data selection

To enrich the dataset for traits exhibiting sex-differential phenotypic coupling that could be decomposed into sex-specific genetic and environmental correlations, we performed a targeted trait-pair selection. We first computed the pairwise phenotypic correlations among 34 UK Biobank traits (6 binary and 28 quantitative) and identified pairs showing the largest male–female differences, ensuring that canonical markers such as standing height, WHR, sex hormones, blood constituents indicative of cardiovascular health, asthma-related markers, selected diseases, and behavioural traits were included. We selected the 250 trait pairs with the strongest sex-difference in phenotypic correlation. Trait abbreviations and sex-specific sample sizes are listed in **Supplementary Table 1**; means and standard deviations (SDs) are provided in **Supplementary Table 2**.

### Phenotypic correlation

Of the 250 trait pairs analysed 15 pairs (6%) exhibited a significant rP only in women, ten pairs (4%) only in men, and 225 pairs (90%) in both sexes. Significance was defined by the 95% confidence interval (CI) excluding zero.

Among the 225 trait pairs with significant rP in both sexes, 68 (30.2%) showed opposite correlation directions, with 33 of these exhibiting a stronger absolute effect in men. The remaining 157 pairs (70.8%) showed same-direction correlations across sexes but with differing magnitudes. In 81 of these (51.6%), the effect was stronger in men (**Figure 2, top left**).

### Sex-specific differences in heritability

Narrow-sense heritability (27) is the proportion of trait variance attributable to additive genetic effects. A difference in heritability between sexes may indicate variation in how much of the trait is explained by genetic factors, suggesting potential sex-specific differences in the underlying genetic architecture (28).

Out of 34 traits, 5 (14.71%) showed nominally significant sex differences in heritability. The largest difference was observed for testosterone (FDR = 1.63e-4) with a higher heritability estimate in men. Educational attainment (Edu), apolipoprotein B (ApoB), C-reactive protein (CRP), and urate also showed nominally significant differences, with higher heritability estimates in women (**Figure 3 left**, **Supplementary Table 4**).

**Figure 3:**
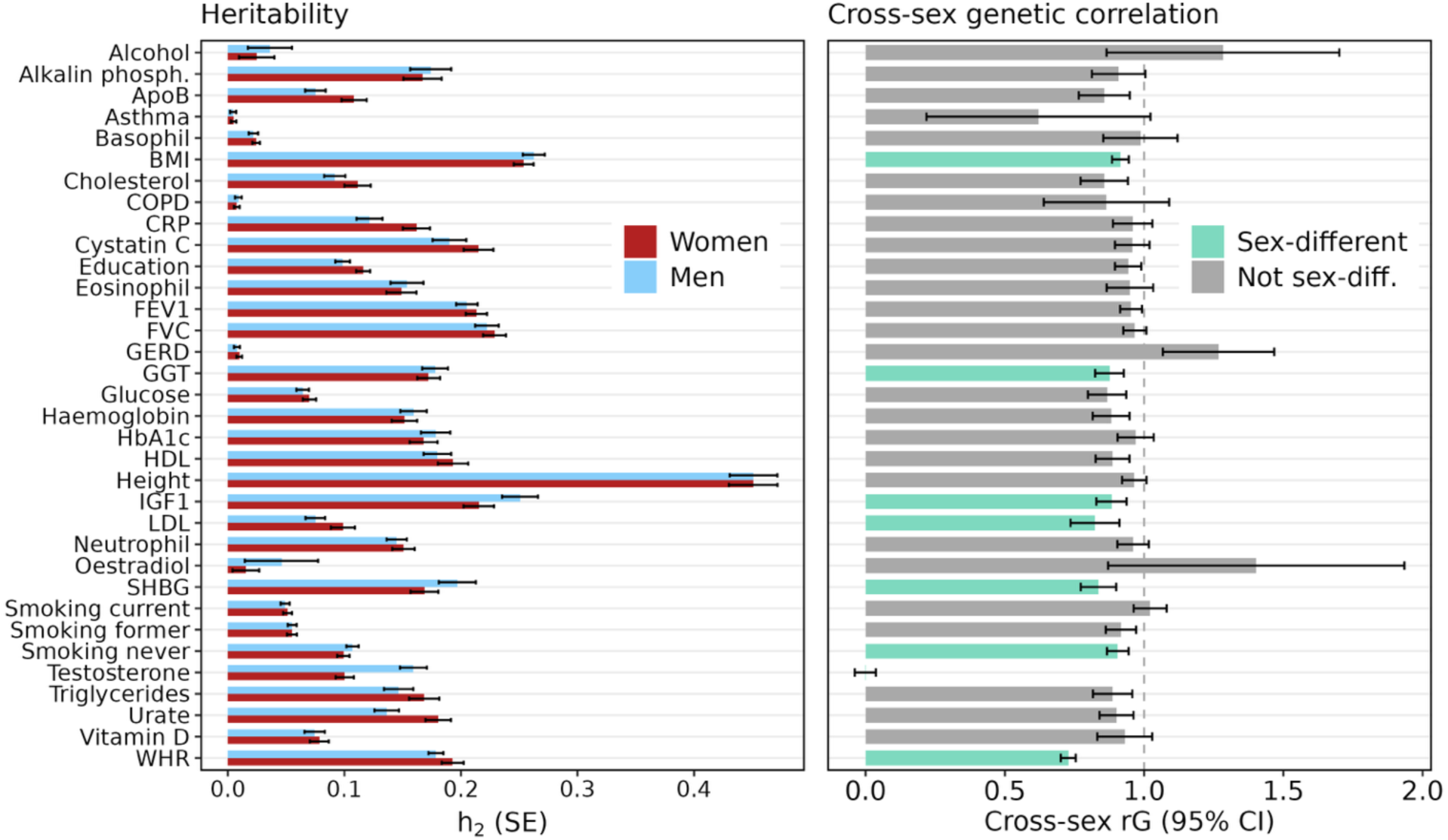
Sex-stratified genetic heritability with standard error (left), Cross-sex (within one trait between men and women) genetic correlation with 95% confidence interval (right). Traits with a genetic correlation significantly different from 1 are coloured in turquoise. This is an indication of sex-different genetic architecture in that trait. After Benjamini-Hochberg correction, only testosterone and WHR remain significantly different from 1, marked with an asterisk. Smoking is abbreviated with Smk.

Since sex-different heritability was observed in only a subset of traits, it is essential to investigate genetic effects more directly, e.g., by estimating genetic correlations between sexes, to better understand these differences.

### Cross-sex same-trait genetic correlation as an indicator of shared genetic architecture between the sexes

Genetic correlations between two population subgroups are commonly interpreted as measures of shared underlying genetic architecture and can estimate the extent of genotype-by-group interactions. Accordingly, genetic correlations between the sexes provide valuable insights into the degree of shared genetic regulation of complex traits and diseases in males and females (12).

We calculated cross-sex rG for all 34 traits, which ranged from -0.0012 (95% CI = -0.08 – 0.07) in testosterone to >1 (1.4 (0.36 – 2.45)) in oestradiol. It should be noted that with very low heritability, LD score regression can yield rG values beyond the theoretically acceptable range of [−1,1] (29).

Of the 34 traits examined, six (17.6%) showed nominal evidence that the cross-sex rG differed significantly from one (**Figure 3**), indicating partial sex-specific genetic architecture. After correction for multiple testing using the FDR, only testosterone and WHR remained significantly below one. These findings are consistent with previous genome-wide studies showing that testosterone levels have largely sex-specific genetic determinants (30) and that WHR exhibits strong sex differences in its genetic architecture (28,31). The deviation of rG from one for testosterone and WHR suggests incomplete overlap in the allelic effects influencing these traits in men and women, pointing to biologically distinct pathways contributing to sexual dimorphism in endocrine and anthropometric traits. Notably, except for testosterone, there was no overlap between traits with a cross-sex rG significantly different from one and those with significantly sex-different heritabilities (**Figure 3**), indicating that differences in the shared genetic correlation between sexes can occur independently of differences in heritability magnitude.

To further understand the sources of sex-different phenotypic correlations between traits, we extended our analysis from within-trait cross-sex genetic correlations to cross-trait correlations. We decomposed sex-different rP into genetic and environmental components by calculating cross-trait genetic and environmental correlations of 299 trait pairs, leveraging phenotypic and genetic correlations along with heritabilities.

### Cross-trait genetic correlation (rG)

Of the 250 trait pairs analysed, 68 (27.2%) showed sex differences in rG (**Figure 2, top right; Figure 4**). Sixteen pairs (23.5%) exhibited significant rG only in women (**Figure 5**).

**Figure 4:**
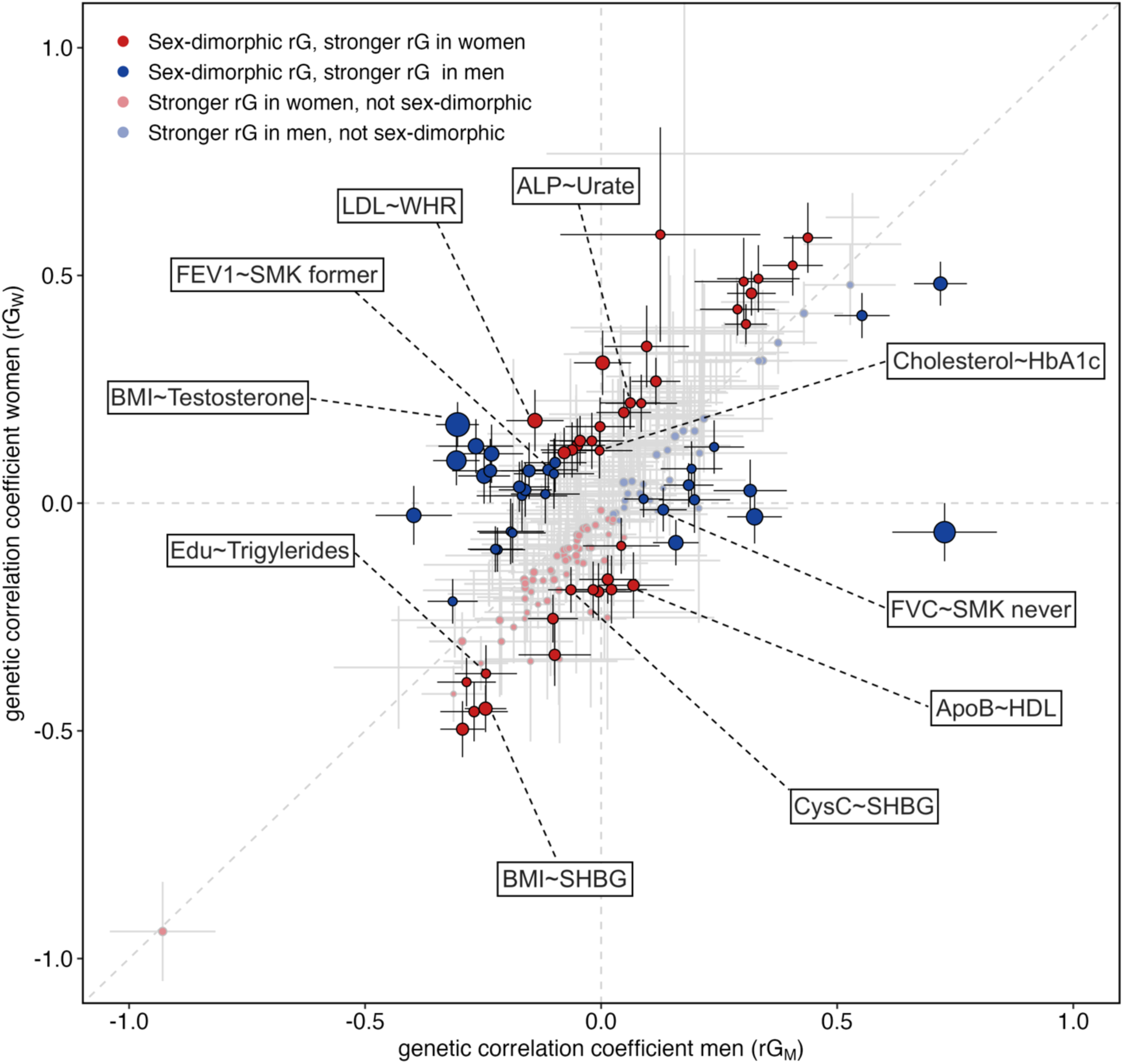
Sex-stratified genetic correlation with 95% confidence interval. Point size according to *p*-value (Benjamini-Hochberg corrected for 250 tests) of *t*-test between men and women, sex-dimorphic trait pairs are shown in bright colour. Red indicates stronger rG in women, blue indicates stronger rG in men. Selected trait pairs showing sex-different genetic correlations are labelled.

**Figure 5:**
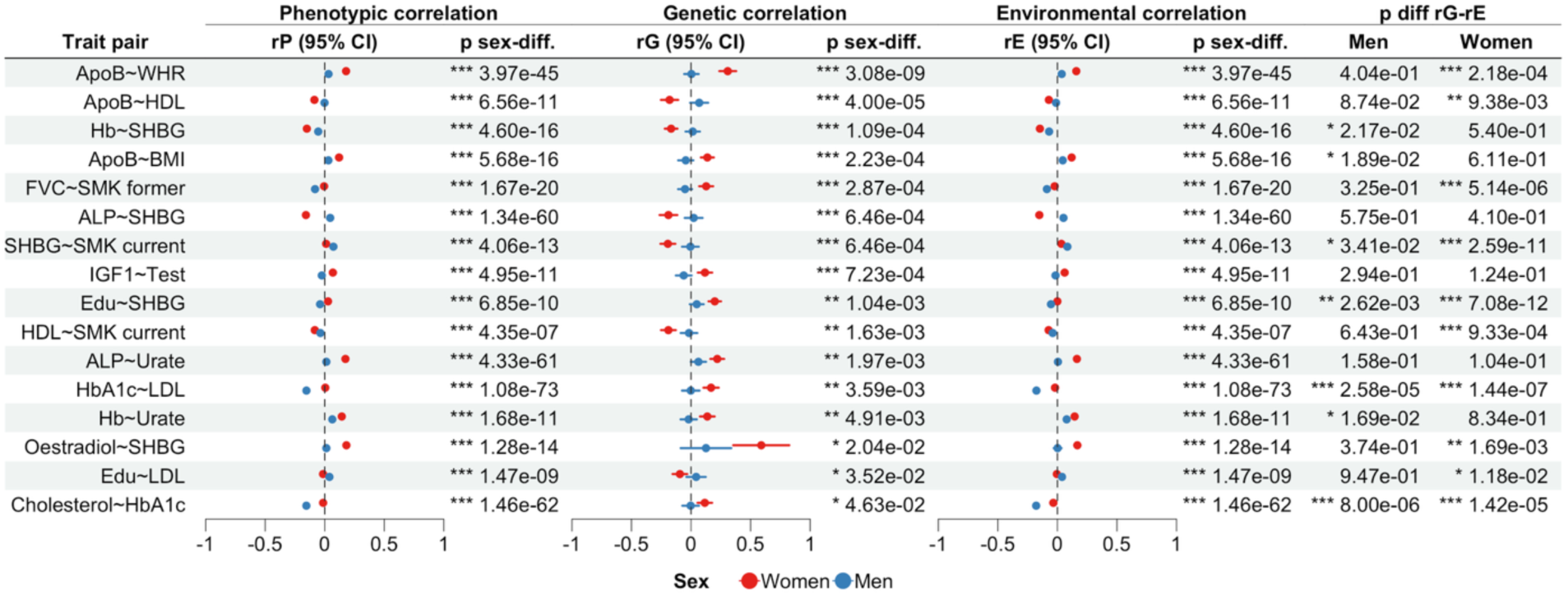
Trait pairs showing significantly sex-different rG having rG only in women. Columns show phenotypic correlation (rP), genetic correlation (rG), and environmental correlation (rE) with 95% confidence intervals (95% CI), their respective *p*-value of the *t*-test between the correlation between men and women (p sex-diff.), and *p*-value of the *t*-test between the rG and rE (p diff rG-rE), separate for men and women.

The identified trait pairs predominantly involved metabolic and behavioral phenotypes, including lipid- and glycaemic-related traits (e.g. ApoB ∼ WHR, ApoB ∼ HDL, HbA1c ∼ LDL, Cholesterol ∼ HbA1c), hormone-related traits (e.g. SHBG with Hb, ALP, smoking behaviour, and oestradiol), and several smoking-associated pairs (FVC ∼ former smoking, HDL ∼ current smoking). Together, these findings indicate sex-specific genetic overlap between lipid metabolism, metabolic regulation, hormonal pathways, and lifestyle-related traits. In all trait pairs sex differences were additionally driven by environmental correlations.

Of the 68 pairs with sex-different genetic correlation 16 pairs (23.5%) showed significant rG only in men (**Figure 6**). The remaining 36 pairs (52.9%) had significant but different rG in both sexes (**Figure 7**). Among these 36 pairs, 25 had rG in the same direction across sexes and 11 had opposite directions; the absolute rG was stronger in men for 16 pairs and stronger in women for 20 pairs.

**Figure 6:**
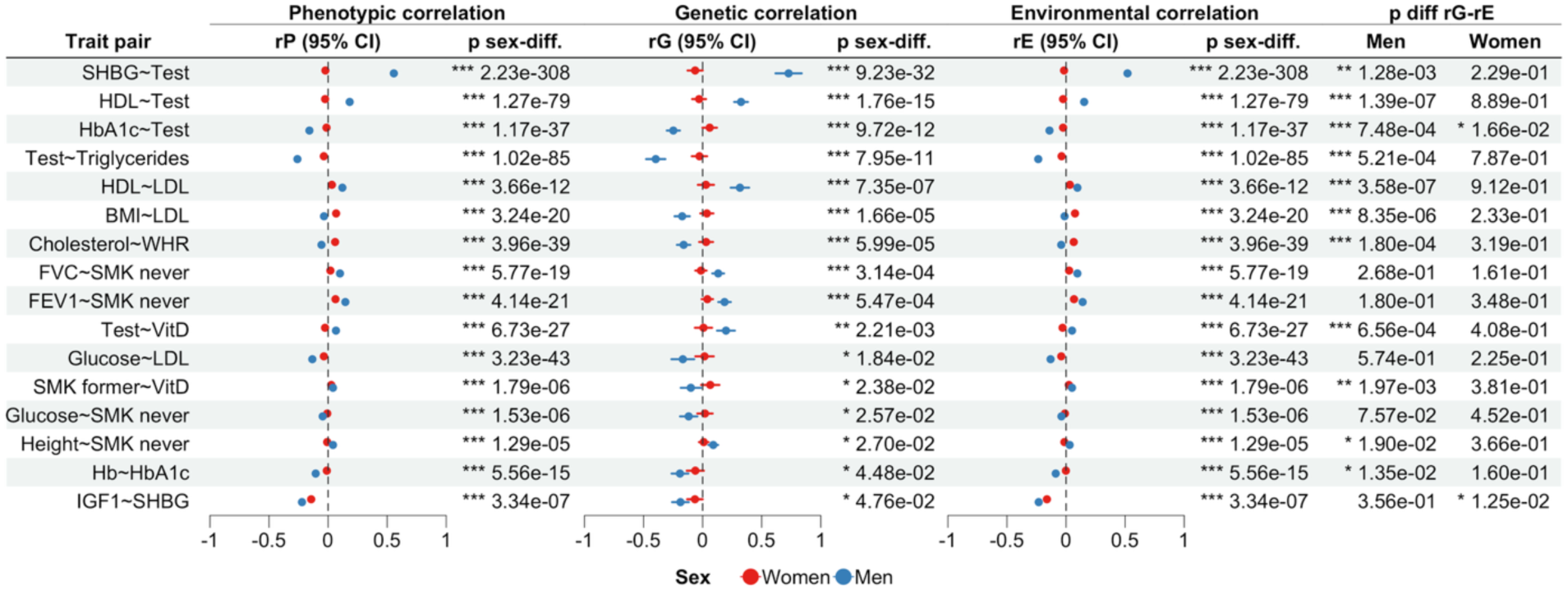
Trait pairs showing significantly sex-different rG having rG only in men. Columns show phenotypic correlation (rP), genetic correlation (rG), and environmental correlation (rE) with 95% confidence intervals (95% CI), their respective *p*-value of the *t*-test between the correlation between men and women (p sex-diff.), and *p*-value of the *t*-test between the rG and rE (p diff rG-rE), separate for men and women.

**Figure 7:**
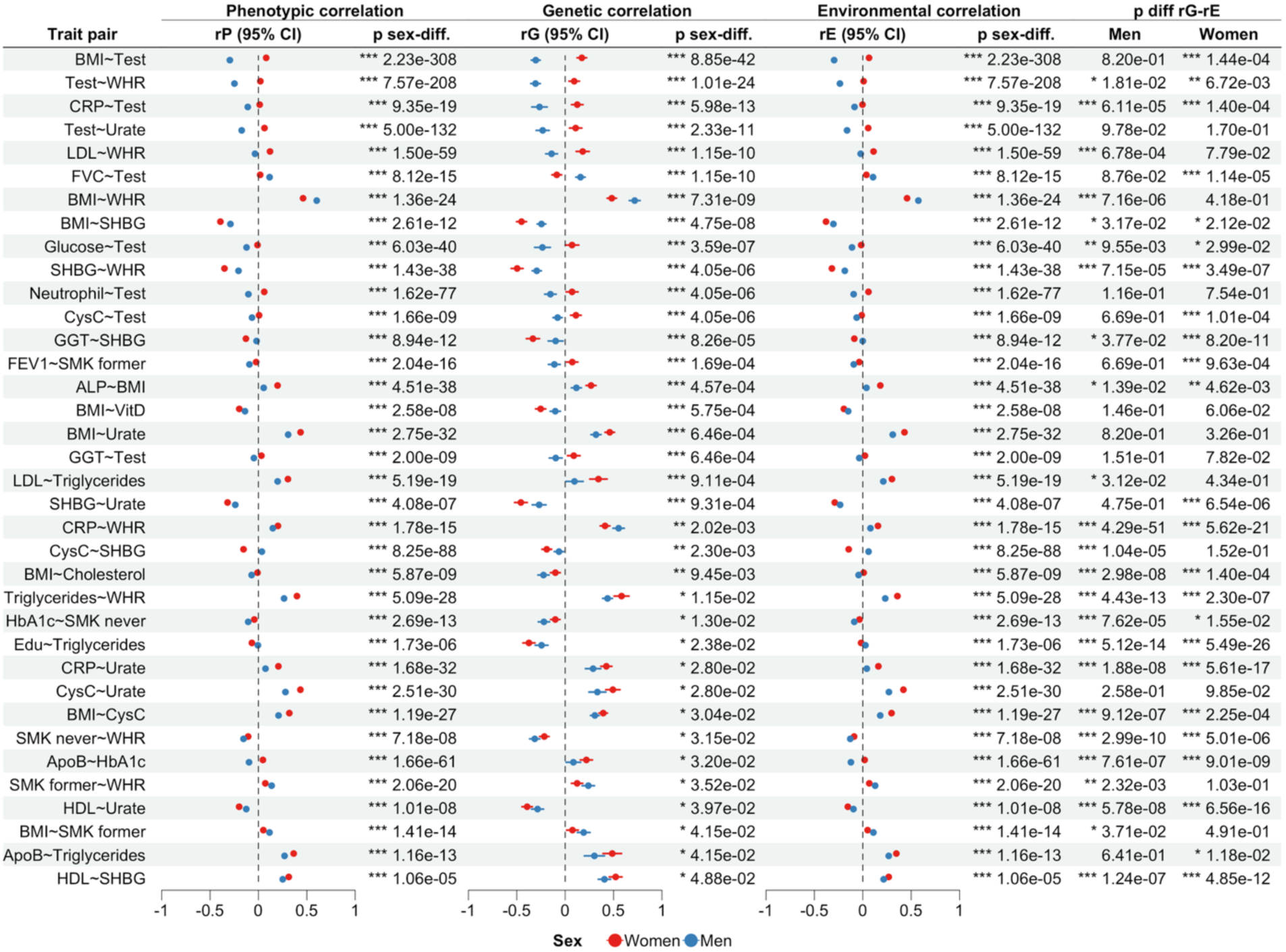
Trait pairs showing significantly sex-different rG with a significant correlation (Significance was defined by the 95% confidence interval excluding zero) in both sexes. Columns show phenotypic correlation (rP), genetic correlation (rG), and environmental correlation (rE) with 95% confidence intervals (95% CI), their respective *p*-value of the *t*-test between the correlation between men and women (p sex-diff.), and *p*-value of the *t*-test between the rG and rE (p diff rG-rE), separate for men and women.

Sixteen pairs (23.5%) showed significant rG only in men (**Figure 6**). The identified pairs predominantly involved endocrine and metabolic traits, including SHBG ∼ testosterone, testosterone ∼ TG, HbA1c ∼ testosterone, and BMI ∼ LDL, indicating male-specific genetic coupling across hormonal regulation and lipid and glucose metabolism. Several associations additionally linked metabolic traits with behavioural or lifestyle factors, such as smoking status with lung function, glucose levels, height, and vitamin D. Together, these results point to male-specific genetic overlap between endocrine regulation, cardiometabolic traits, and lifestyle-related exposures.

Among the 36 pairs significant in both sexes (**Figure 7**), the majority involved metabolic, anthropometric, and lipid-related traits, including BMI, WHR, lipid fractions, glycemic markers, and inflammatory biomarkers (e.g. BMI ∼ WHR, LDL ∼ WHR, BMI ∼ urate, SHBG ∼ WHR, ApoB ∼ HbA1c). Several pairs additionally linked metabolic traits with hormonal or renal markers, such as testosterone, SHBG, cystatin C, and urate. Overall, these results indicate shared genetic architecture between men and women across cardiometabolic and body-composition traits, with sex-specific differences in effect magnitude and direction.

### Top traits sex-different rG

Of the top 10 trait pairs showing the strongest sex differences in genetic correlation, nine involve testosterone and are associated with BMI, SHBG, WHR, HDL, CRP, HbA1c, urate, triglycerides, and FVC, in addition to the LDL ∼ WHR pair (**Supplementary Figure 1**).

BMI with SHBG (rG_m_ = -0.3, rG_f_ = 0.17) shows the most significant sex-difference in rG. All of these trait pairs also showed significant sex differences in environmental correlation (rE). Additionally, all mentioned pairs except for testosterone with BMI and urate as well as LDL ∼ WHR had a significant difference between rG and rE in men, while only five of the ten pairs (testosterone with BMI, urate, CRP, HbA1c and with FVC) did so in women.

### Traits with high uncertainty in rG

We observed that the traits oestradiol, alcohol, and the diseases asthma, COPD and GERD show great uncertainty in their rG calculation. For oestradiol and alcohol, this might stem from very small sample sizes (oestradiol (N = 51,218), alcohol (N = 146,072), see **Supplementary Table 1**). For asthma, COPD and GERD, this might be due to smaller sample sizes in combination with them being binary traits with lower case numbers (asthma (N = 267,450), COPD (N = 290,753) and GERD (N = 283,329), see **Supplementary Table 1**). **Environmental correlation**

All 250 trait pairs showed significant sex differences in environmental correlation (rE) (**Figure 2, bottom left** and **Figure 8**). Twentytwo pairs exhibited significant rE only in men, and 24 pairs only in women. The majority of trait pairs – 204 – had significant rE in both sexes. Of these, 158 pairs showed rE in the same direction in men and women, with 92 (45.1 %) having stronger rE in women. The remaining 54 pairs had rE in opposite directions across the sexes, and in most of these (41) the absolute effect was stronger in men.

**Figure 8:**
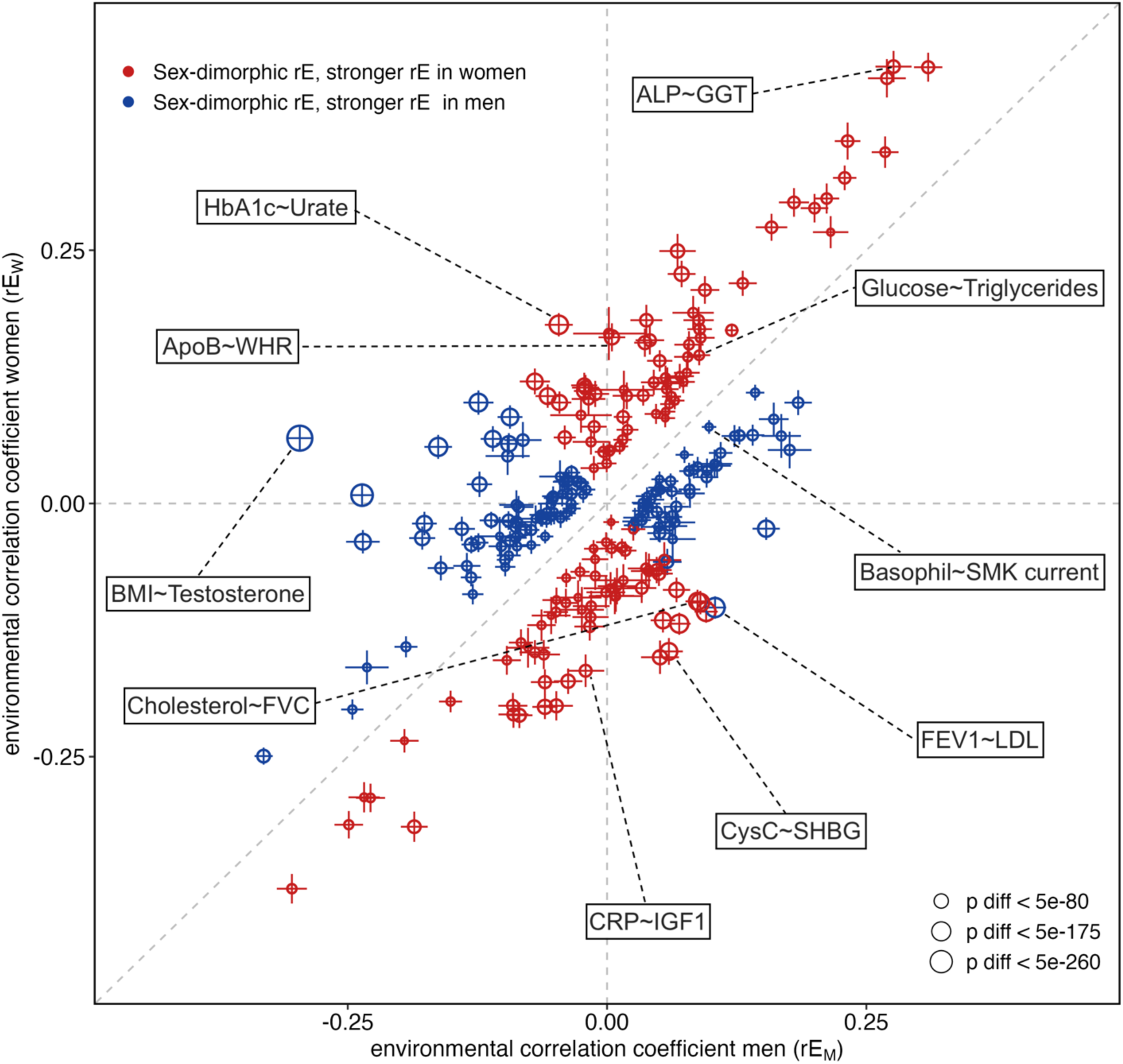
Sex-stratified environmental correlation with 95% confidence interval. Point size according to *p*-value (Benjamini-Hochberg corrected for 250 tests) of *t*-test between men and women. Selected trait pairs showing sex-different environmental correlations are labelled.

### Top traits sex-different rE

Trait pairs showing the strongest sex differences in rE (top 10) include testosterone ∼ SHGB, which exhibited the largest difference (rEm = 0.52, rEf = -0.02), as well as testosterone with BMI, WHR and urate. Additional pairs involved lung function measures in combination with lipid traits (LDL, cholesterol, SHBG, and ApoB), and the HbA1c ∼ urate pair (**Supplementary Figure 2**). The traits correlated to testosterone additionally show sex-different rG indicating sex-different genetic components; six pairs show a significant difference between rG and rE in men (testosterone with SHBG and with WHR, HbA1c ∼ urate, and lung function measurements with LDL, SHBG and ApoB) and five in women (testosterone with BMI and with WHR, lung measurements with LDL and cholesterol and HbA1c ∼ urate).

### Sex-different rE as primary contributor to sex-different rP

The majority (182, 72.8%) of the sex-difference in phenotypic correlation could be attributed solely to sex-different environmental correlation (rE). An additional 68 pairs (27.2%) exhibited sex differences in both environmental (rE) and genetic (rG) correlation. No pairs were affected exclusively by sex-specific rG.

Overall, our findings indicate that sex differences in phenotypic correlations are more frequently driven by environmental rather than genetic factors. However, this pattern may also reflect lower statistical precision in the estimation of genetic correlations.

### Frequency of traits being involved in sex-different rP, rG and rE

**Supplementary Table 6** summarizes how frequently individual traits appear in pairs exhibiting significant sex differences in phenotypic, genetic, and environmental correlations. Overall, traits related to endocrine function, metabolism, and cardiometabolic health were most frequently involved. SHBG and testosterone were the most prominent traits, each appearing in 25 trait pairs with significant sex differences at the phenotypic and environmental levels, and in 13 and 15 pairs, respectively, at the genetic level. This highlights the central role of sex hormones in driving sex-specific correlation patterns across a wide range of traits.

Several metabolic and cardiometabolic traits also appeared frequently, including CysC, urate, ALP, ApoB, Hb LDL, TG, cholesterol, BMI, and smoking status (never). These traits were commonly involved in sex-differentiated phenotypic and environmental correlations, whereas comparatively fewer showed significant sex differences at the genetic level, with BMI being a notable exception. This pattern suggests that non-genetic factors contribute substantially to the observed sex-specific relationships.

In contrast, genetic sex differences were concentrated in a smaller subset of traits, including SHBG, testosterone, WHR, BMI, and selected lipid and glycaemic traits. Several traits including GERD, alcohol consumption, COPD, asthma, eosinophil and basophil count showed no significant sex differences in genetic correlation despite appearing in multiple phenotypic or environmental associations. This pattern indicates that for many traits, sex differences in phenotypic correlations arise primarily from environmental variation rather than from underlying genetic architecture.

Taken together, these results suggest that phenotypic sex differences in trait–trait correlations are widespread but are predominantly driven by environmental components. Genetic sex differences appear more restricted and are largely confined to traits related to hormonal regulation, metabolism, and body composition, underscoring the importance of endocrine pathways in shaping sex-specific genetic architectures.

### Traits correlated with smoking

Among the investigated trait pairs, 44 involved a smoking phenotype (never, former, or current). All of these exhibited significant sex differences in environmental correlation, whereas only 13 showed sex differences in genetic correlation. Of those trait pairs the strongest sex-differences in rG have been observe in traits correlated to lung function (FEV1 and FVC).

Notably, for the trait pair CRP and non-smoking, there is no evidence of sex differences in rE, indicating that the sex differences in rP in these traits are primarily driven by rG.

### Testosterone

Testosterone shows significantly different heritability across the sexes after multiple testing correction (h^2^_m_= 0.16; 95% CI = 0.14 – 0.18; h^2^_f_= 0.1; 95% CI = 0.09 – 0.12) and a cross-sex rG being significantly different from 1 after multiple testing correction (cross-sex rG = -0.001, 95% CI = -0.08 – 0.07). We tested testosterone in 25 trait pairs for sex-different genetic and environmental correlations. All 25 pairs show sex-different environmental correlations (**Supplementary Figure 3**).

In contrast, 15 of the 25 trait pairs also showed significant sex differences in genetic correlation, indicating that genetic effects contribute to sex-specific coupling for a subset of testosterone-associated traits. These included metabolic, inflammatory, and anthropometric traits such as BMI, WHR, triglycerides, HDL, HbA1c, CRP, urate, and lung function measures. Several associations, including testosterone with BMI, WHR, and lipid traits, displayed pronounced divergence between men and women with most of them having opposing effects or only an effect in one sex.

Together, these findings indicate that while testosterone-related trait associations are universally influenced by sex-specific environmental factors, only a subset is driven by sex-differentiated genetic architecture. This highlights testosterone as a central node in sex-specific regulation of metabolic, cardiometabolic, and inflammatory traits, with environmental influences playing a dominant role across most trait pairs.

### Genetic versus environmental correlation

According to the gene-environment equivalence assumption of Mendelian Randomisation (MR), the genetic and environmental correlations between two traits are expected to agree if a phenotypic correlation is due to (bidirectional) causal effects. If they differ, it may indicate the presence of confounders of a different nature: e.g. strong environmental confounders lead to dominant environmental correlation (32,33).

We found 87 trait pairs with agreeing rG and rE in both men and women. Among those, 17 trait pairs show agreeing rG and rE only in women, and 5 trait pairs show agreeing rG and rE only in men, indicating potentially bidirectional causal effects in those trait pairs (**Figure 9**).

**Figure 9:**
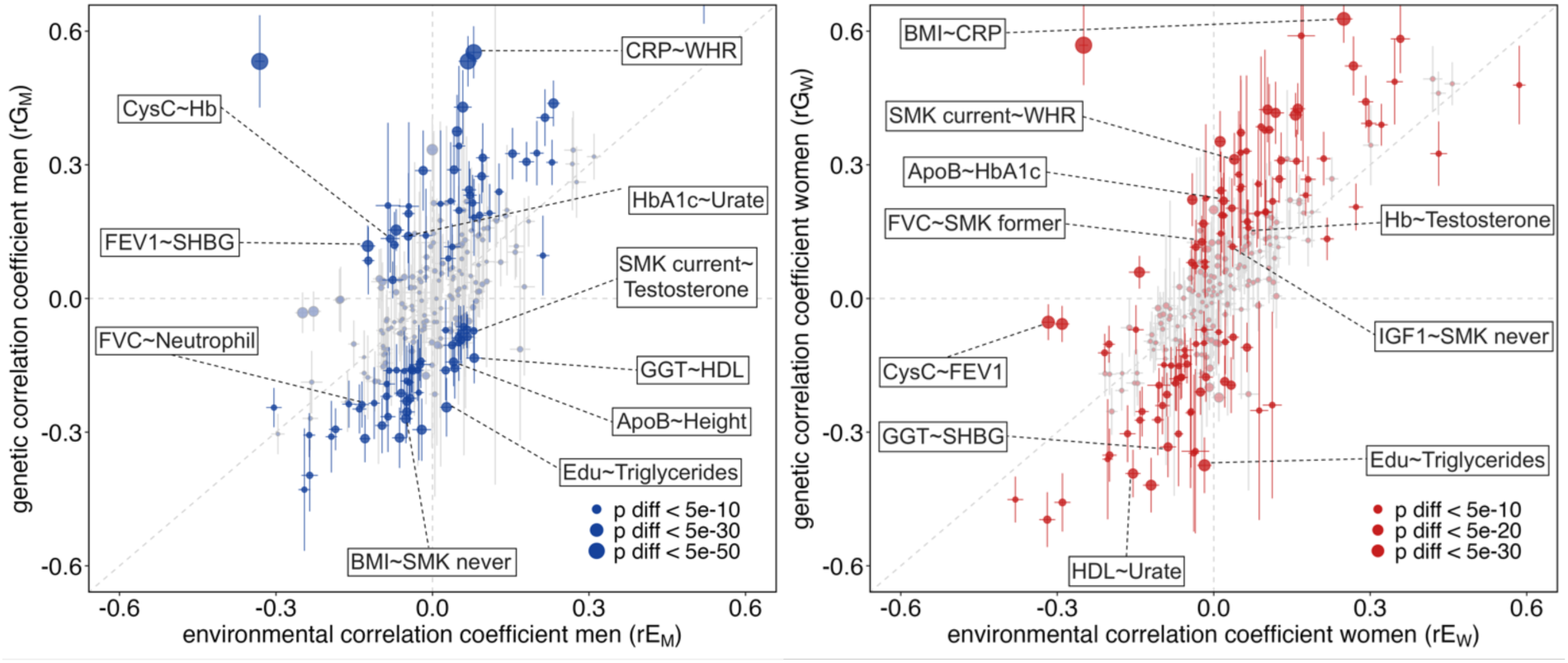
Sex-stratified genetic correlation (rG) versus environmental correlation (rE) with 95% confidence intervals (left: men, right: women). Trait pairs with significant rG and rE, and a significant difference between rG and rE in the respective sex, are brightly coloured. Dot size reflects the significance of the difference between rG and rE. Selected trait pairs showing significant differences between genetic and environmental correlations in the respective sex are labelled.

On the other hand, **Figure** 10 shows the top 5 trait pairs with disagreeing rG and rE in both sexes, men only and women only. It so worth noting that for men 4 out of the top 5 traits with disagreeing genetic and environmental correlation are correlated to lung function.

**Figure 10:**
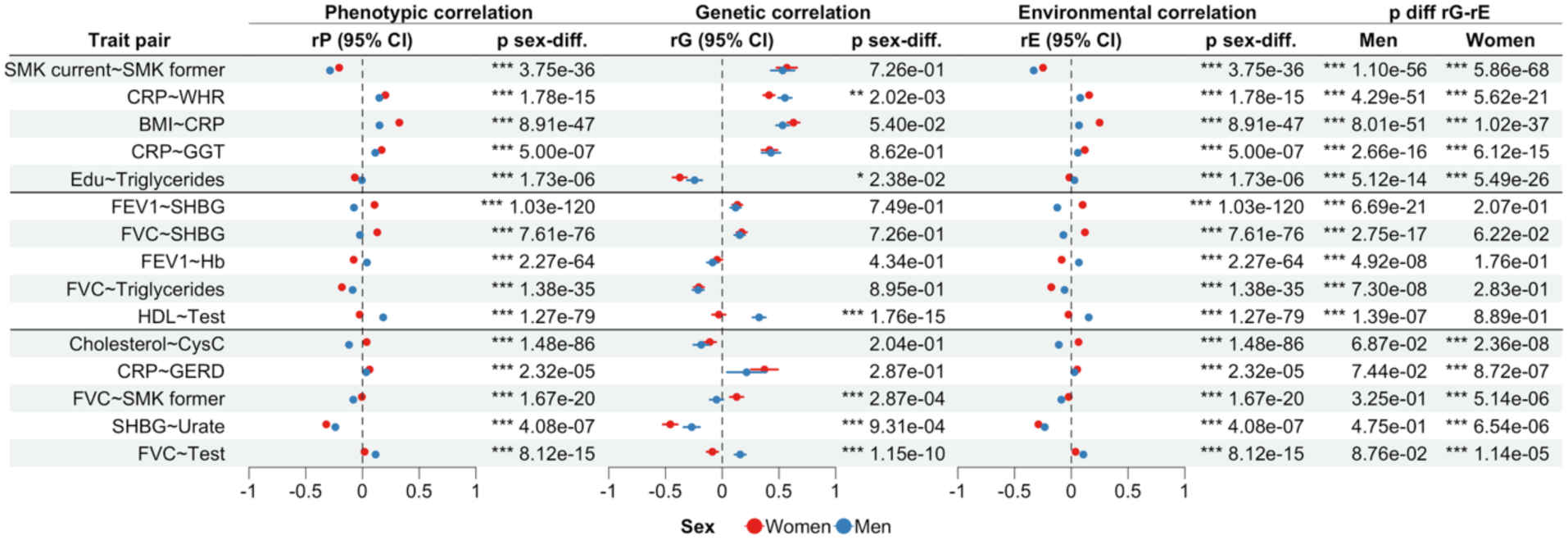
Showing top 5 traits with disagreeing rG and rE in both sexes (top), only in men (middle) and only in women (bottom). Columns show phenotypic correlation (rP), genetic correlation (rG), and environmental correlation (rE) with 95% confidence intervals (95% CI), *p*-value (*t*-test) of sex-difference in the respective correlation (p sex-diff.) determined by *t*-test, and *p*-value of difference between rG and rE (p diff rG-rE) determined by *t*-test in men and women.

Additionally, those traits show sex-different environmental correlation and no sex-difference in genetic correlation. This is an indicator of potential sex-differences in environmental correlations or sex-specific environmental confounders lead to the sex-different phenotypic correlations.

For women, trait pairs with significantly different genetic and environmental correlations, traits span biomarkers of lung function, cardiometabolic health, renal function, systemic inflammation, endocrine regulation, gastrointestinal disease, and lifestyle exposures.

## Discussion

We investigated sex-different genetic and environmental contributions to sex-different rP of 250 trait pairs from the UK Biobank. The trait pairs were selected according to sex-different rP while including basic phenotypic measurements. The trait pairs consist of 34 different traits. To assess the genetic contribution to those sex differences in the 34 traits, we calculated, as a first step, the sex-stratified heritability Calculating the sex-stratified heritabilities and comparing them across the sexes provides information about varying genetic contributions across the sexes. We found nominally significant sex-different heritability for five traits (Edu, ApoB, CRP, urate, and testosterone). After multiple testing corrections, only testosterone remained significant. The sex-different heritabilities in those five traits contribute to explaining the sexually dimorphic phenotypes. Men show up to 15 times higher testosterone levels compared to women (34). The sex-different heritability is reflected in the sex-different phenotype. Further, studies show that females perform better during school education (35). Differences in heritability do not necessarily indicate differences in genetic architecture, as they may result from variations in environmental variances between the sexes, as also shown by Rawlik *et al.* (12).

Interestingly, the traits such as height and SHBG, which are known for sex-different phenotypes (36,37), do not show significant differences in heritability between sexes, which might indicate non-heritable sources for the sex-different phenotypes.

Traits with cross-sex rG significantly different from one and that frequently exhibit sex-different genetic correlations with other traits provide strong evidence for a sex-specific genetic architecture.

Testosterone and WHR show sex differences in their phenotypes, and we can find a sex-different genetic contribution by finding a cross-sex rG different from one after multiple testing correction. The genetic architecture of WHR has previously been found to be sex-different. GWASs have repeatedly found more genetic associations of WHR in women than men (31,38).

There is no overlap between traits showing sex-different heritabilities and a cross-sex rG different from 1, except for testosterone, indicating that genetic architectures may be discrepant, but this does not necessarily lead to different narrow-sense heritability.

To assess the sex-different environmental contribution to sex-different phenotypes, we calculated the sex-stratified rE. For that, we used the previously calculated rP, rG and heritabilities, decomposing rP into sex-specific genetic and environmental contributions.

In this study, we aimed to investigate whether observed sex differences in phenotypic correlations could be attributed to sex-specific genetic effects, environmental influences, or a combination of both. We can see that while the strength of genetic or environmental contributions is specific to each trait pair, an overall trend of stronger sex differences in the environmental components is observed. However, this insight should be treated with caution. Genetic correlations are estimated with larger errors, which reduces power to detect sex-differences and hence could lead to an underestimation. Sex-differentiated genetic and environmental effects have been investigated previously, with substantial evidence for either type, supporting the existence of sex-specific genetic and environmental effects (12,39,40).

Disagreeing rG and rE may be indicative of potential non-genetic confounders. Specifically for smoking, previous studies have found that professional occupation and, with that, social strata act as a non-heritable confounder to smoking (41).

Non-heritable confounders can also include population stratification. A study has shown that geographic location influences the association between genetic variants and complex traits (42). Population stratification can partially be corrected for by using the first 10 principal components as covariates; however, it is not sufficient to eliminate all confounding. Additional sources of environmental confounding include indirect genetic effects. Indirect genetic effects are effects of alleles in parents or siblings that are mediated through the environment created by relatives who have the respective allele (43,44).

Testosterone is one of the repeatedly appearing biomarkers regarding sex-specific differences in correlation in this study. Testosterone can appear in two forms in the blood: bound and unbound. Most of the total testosterone is bound to blood plasma proteins like SHBG. The unbound or free testosterone, typically 2-3%, is thought to have androgenic effects. Testosterone can weakly bind to albumin and retain its androgenic effects. When bound to SHBG, testosterone loses its effects and becomes inactive (45). As men have up to 15 times higher testosterone levels than women of the same age (34), it is unsurprising to find differing phenotypic correlation between testosterone and SHBG (t_rP_ = 160.59; FDR = < 2.23e-308) having strong rP in men (rP_m_ = 0.56, 95% CI = 0.552 – 0.56) while having a negligible rP in women (rP_f_ = -0.02, 95% CI = -0.029 – -0.017). Elevated testosterone levels are shown to have differing effects on men and women with regard to different diseases (30). Similarly to phenotypic correlation, the trait pair testosterone ∼ SHBG shows strong genetic correlation in males (rG_m_ = 0.73, 95% CI = 0.62 – 0.84) and negligible correlation in females (rG_f_ = -0.06, 95% CI = -0.13 – 0.0002) (t_rG_ = 12.13, FDR = 9.23e-32) which is confirming previous findings (46). We further found that testosterone and SHBG demonstrate environmental correlation in men (rE_m_ = 0.52, 95% CI = 0.5 – 0.54) but not in women (rE_f_ = -0.017, 95% CI = -0.029 – -0.005) (t_rE_= 38.55, FDR = < 2.23e-308).

Testosterone also shows sex-different correlation with WHR, urate, and TG. Urate is the end product of purine metabolism in humans, and increased levels have been associated with inflammatory effects (47). Men show higher circulating urate levels than women (**Supplementary Table 2**). Sex-different urate levels have been found to correlate with differing testosterone and SHBG levels (48). Similarly, we can observe that rG and rE between urate and testosterone and urate with SHBG differ significantly for men and women. As identified by the epigraph database using MR (49) potential confounders for urate ∼ testosterone include TG, hip circumference and ApoB (*p* < 1e-3). ApoB additionally shows a nominally sex-different heritability, which could be a contributing factor to the sex-different correlations. Rasheed *et al.* (50) have found increased very low-density lipoprotein TG to be associated with the risk of gout, suggesting pleiotropic effects of ApoB in determining gout. ApoB is present on very low-density lipoprotein TG, and gout is a disease in which urate crystals deposit in joints (50).

Similarly, the epigraph database suggests 278 potential confounders (*p* < 1e-3) using MR (49) for the trait pair urate ∼ SHBG, which include TG, BMI, and HDL.

### BMI ∼ CRP

Looking at specific trait pairs, BMI shows differing sex-specific phenotypic correlation with CRP (rP_m_ = 0.15, 95% CI = 0.14 – 0.16; rP_f_ = 0.324, 95% CI = 0.320 – 0.328; t_rP_ = -48.26, FDR = < 2.23e-308). CRP is a phase protein that rises in response to inflammation and is used as a biomarker for early detection of infection in the emergency room (51). However, at low concentrations, tests are not specific, making it difficult to locate and identify the exact inflammation (51). We can confirm previous findings that the pair BMI and CRP shows a stronger rP in women than in men (52).

The trait pair shows genetic correlation; however, the sex-specific difference is not significant (rG_m_ = 0.53, 95% CI = 0.48 – 0.59; rG_f_ = 0.63, 95% CI = 0.57 – 0.68; t_rG_ = - 2.39, FDR = 5.40e-02). The environmental correlation shows a significant sex-specific difference with low environmental correlation in males (rE_m_ = 0.07, 95% CI = 0.05 – 0.08) and higher environmental correlation in females (rE_f_ = 0.25, 95% CI = 0.23 – 0.27; t_rE_ = -14.35, FDR = 1.10e-49).

Our results show that the stronger phenotypic correlation in females in the trait pair BMI-CRP can be explained to a greater extent by a strong environmental correlation of the two traits rather than a genetic correlation. This could point to the theory that elevated BMI levels in females and with that elevated CRP levels stem to a greater extent from external impacts in females compared to men. Such external impacts could be attributed to lifestyle and behaviour, such as the amount of physical activity. There are studies showing that women engage less in physical activity compared to men (53,54).

### HbA1c ∼ LDL

HbA1c is a form of Hb that is linked to sugar (55). It is used as a marker for identifying diabetes as it reflects average glucose levels of the last eight to twelve weeks (56).

LDL is considered the “bad” cholesterol. It is responsible for transporting cholesterol to the cells (57). However, too high LDL levels can cause a buildup in the artery walls and increase the risk of coronary artery disease (58) or a stroke (59).

Statins is a medication used for lowering cholesterol levels, especially for LDL-cholesterol and therefore lowering the risk of cardiovascular disease (60–62). As a side effect it increases the risk of diabetes (63–65). In men, we observe a negative phenotypic correlation (rP_m_ = -0.15, 95% CI = -0.16 – -0.148) and negative environmental correlation (rE_m_ = - 0.18, 95% CI = -0.19 – -0.16) between LDL and HbA1c.

The described side effect of statins has been observed to be stronger in women (66–68) which would lead to a stronger negative phenotypic correlation in women. However, we observe no significant phenotypic or environmental correlation in women and only a genetic correlation (rG_f_ = 0.17, 95% CI = 0.1 – 0.23). This inconsistency indicates the potential presence of a genetic confounder.

### Cholesterol ∼ CysC

Cholesterol and CysC have disagreeing rG and rE only in women, which is indicative of a non-heritable confounder affecting this trait pair only in women. The trait pair shows sex-different rP and rE with a stronger negative correlation in men and a weaker positive correlation in women. There is no sex-different rG. This is indicative that sex-different environmental components or confounders contribute to the sex-different rP rather than sex-different genetics.

CysC is a marker of renal function and kidney disease (69) and cardiovascular risk (70). Cholesterol comprises total cholesterol levels of high- and low-density lipoprotein (71). Werner *et al.* found a sex-specific effect of age on CysC levels (72) which could be indicative of sex-different mechanisms governing CysC levels. Additionally, smoking could be a non-heritable confounder that affects both CysC and cholesterol. Smoking influences levels of both total cholesterol and LDL (73,74) while there is evidence for and against the influence of smoking on CysC (75,76).

As (total) cholesterol is a composite measure of both HDL and LDL, it would be valuable to investigate HDL and LDL levels separately.

### FEV1 ∼ Hb

FEV1 quantifies the volume of air exhaled during the first second of a forced expiratory breath and serves as a key indicator of the severity of obstructive pulmonary diseases, including COPD and asthma (77). Consistent with known physiological differences, men exhibit higher mean FEV1 values than women (**Supplementary Table 2**).

Hemoglobin (Hb) is the oxygen-transporting protein in erythrocytes (78), with higher average concentrations observed in men (79) (**Supplementary Table 2**).

The FEV1–Hb trait pair exhibits a discrepancy between rG and rE, indicating the presence of potential non-genetic confounding factors. Moreover, the results suggest sex-specific differences in rP and rE, but not in rG. Notably, FEV1 and Hb show opposing rP and rE across sexes, displaying a negative correlation in women and a positive correlation in men, whereas rG is negative in both sexes.

### CRP ∼ Smoking

CRP is a protein produced by the liver in response to inflammation. It is part of the acute phase response and is released into the bloodstream when there is tissue injury, infection, or inflammation. CRP is widely used as a biomarker for systemic inflammation and can be measured through a simple blood test (84). Elevated CRP levels are associated with a wide range of conditions, including infections, autoimmune disorders, cardiovascular disease, and certain cancers (85,86). However, it is a nonspecific marker and should be interpreted in the context of other clinical findings (87).

Literature suggests an inconsistency in findings about the association of smoking and CRP levels and also a variation by sex (88,89). In an early study, Das (90) reported significantly higher CRP levels in smokers compared to non-smokers, with higher CRP levels in female non-smokers compared to male non-smokers (median values of 1.0 mg/l and 11.2 mg/l for male non-smokers and smokers, respectively, and for females 2.0 mg/l and 11.6 mg/l, respectively). In a more recent study, Koh *et al.* (89) detected a male-specific association between urine cotinine and CRP in male smokers that was not detected in female smokers. Upon smoking cessation, CRP levels fail to decrease immediately, which points to a recovery period of the tissue damaged by smoking (91,92).

We find a positive sex-different rP for current smokers with a significantly stronger association in men (rP_m_ = 0.07, 95% CI = 0.07 – 0.08; rP_f_ = 0.04, 95% CI = 0.04 – 0.05; FDR = 2.56e-07). A sex-specific increase of CRP levels with smoking only in males has been observed previously (89).

For current smokers, the sex-different rP can be more attributed to sex-different rE with rE in men but not in women (rE_m_ = 0.05, 95% CI = 0.04 – 0.06; rE_f_ = 0.01, 95% CI = 0 – 0.02; FDR = 2.56e-07) and no sex-different rG (rG_m_ = 0.38, 95% CI = 0.29 - 0.46; rG_f_ = 0.35, 95% CI = 0.28 - 0.42; FDR = 0.77).

This points to the fact that the sex differences between smokers and CRP in rP can primarily be accounted for by sex-different environmental effects. For all of 44 pairs involving a smoking phenotype, significant sex-differences in environmental correlation can be observed. Only 13 out of 44 pairs additionally show sex-different genetic correlation. Those trait pairs include smoking and lung function, BMI, SHBG, HbA1c, WHR, HDL, Glucose, VitD and height.

### Conclusion and limitations

In conclusion, we investigated 250 trait pairs to identify environmental or genetic causes for sex-specific discrepancies in the phenotypic correlation of those trait pairs. We confirmed trait-specific previous findings of sex-specific differences in phenotypic correlation and investigated some of those trait pairs in more detail to explain whether those sex-specific discrepancies stem from sex-specific genetic or environmental effects. We found that, to a greater extent, environmental mechanisms are responsible for the sex-specific differences in phenotypic correlation.

Using the large sample size available in the UK Biobank enabled the accurate estimation of nuanced differences on an unprecedented scale. Despite the large sample size, results might not be transferable to other populations. While a high educational level, for example, is generally associated with lower cholesterol levels in developed countries, an inverse relationship of higher cholesterol levels in the group of the highest socio-economic stratum can be seen in developing countries (93). This is an indicator of different lifestyles in higher-income societal classes between developed and developing countries. Limited availability of genetic data for, for example, African (94) countries additionally restricts the possibilities to conduct similar large-scale genetic analysis in populations of other ancestries. Therefore, a limitation of this study is that results are very specific to the UK population and within that population, middle- to high-income classes might be overrepresented (95).

Further distinctions should be made when analysing data from women, as their pre- or post-menopausal status, as well as hormonal contraceptive use, can influence biomarker levels (96,97). One study found a different genetic basis for pre- and post-menopausal women for BMI, waist, and hip circumferences. Findings show a lower h² for waist circumference and WHR in pre-menopausal women, which suggests a greater environmental impact on hip circumference in this group (96). In a study on biomarkers by Ramsey *et al.* (97) a simulation reported up to 41% false discoveries when pre-menopausal women were not corrected for hormonal contraceptive use. Hormonal contraceptive users exhibited significantly higher CRP levels, along with correlation with the biomarkers interleukin-6 and soluble tumour necrosis factor, which was not observed in non-contraceptive users (98). Moreover, CRP levels also vary according to the phases of the menstrual cycle in both contraceptive (97) and non-contraceptive users, with nearly twice as many women showing elevated CRP levels (> 3 mg/L), passing the threshold of an elevated risk of cardiovascular disease, and cholesterol levels (≥200 mg/dL) warranting therapy (99). In particular, oestradiol requires cycle phase-specific measurement (100). Given the evidence, categorising women into pre- and post-menopausal groups, and further into menstrual phases, is necessary and would substantially enhance the precision of biomarker correlations. However, in this dataset, results are already affected when the sample size falls below 300,000, and further stratification by subgrouping women could introduce significant uncertainty. Nonetheless, whenever possible, women should be stratified by menopausal and menstrual status at the time of measurement, as well as by hormonal contraception use.

Additionally, CRP levels respond to acute inflammation or infection with an increase of up to and exceeding 500 mg/L. The half-life of the protein in circulation is 19 hours. It is possible that CRP levels could remain elevated for up to a week following the initial increase. A serial measurement instead of single time points would potentially yield a more accurate reflection of CRP levels (92,101). The measurements in this study were conducted at two time points, 2 to 6 years apart. A more frequent serial measurement over a period of several weeks would improve the accuracy, and it would be more robust to peaks due to mild inflammations such as sporadic acute illness.

Further, in certain trait pairs, where there is a non-linear relationship between two traits and men and women have markedly different levels of one of these traits, the difference in correlation may be sex-unrelated. This could be tested in future studies using non-linear models connecting the two traits.

In future research, we aim to employ sex-stratified Mendelian Randomisation to meticulously examine these 34 traits, to discern the directional causality of the observed phenotypic correlations in both sexes. This will involve a detailed analysis framework that accounts for potential pleiotropy and other confounders, thereby providing robust insights into the genetic underpinnings of sex differences in health and disease.

Future research could aim at systematically identifying potential confounders of trait pairs, for example, BMI is a confounder of the trait pair testosterone and SHBG (49). Hence, it remains to be examined if the correlation between testosterone and SHBG can potentially be explained by the confounder of BMI. That could clarify whether the sex differences in phenotypic correlations are due to genetic differences alone or whether they are also influenced by environmental factors such as lifestyle, which is often reflected in BMI. Thus, in general, future work could examine systematically what role confounders play in the observed sexual dimorphisms of trait correlations.

## Data Availability

The data used in this study are from the UK Biobank and cannot be shared publicly by the authors owing to participant-consent and data-governance restrictions. UK Biobank data are available to bona fide researchers upon application via the UK Biobank Access Management System (https://www.ukbiobank.ac.uk). This research was conducted under UK Biobank application number 16389. The genome-wide association summary statistics used for 33 of the 34 traits are publicly available from the Neale Lab (http://www.nealelab.is/uk-biobank); summary statistics for waist-to-hip ratio were generated by the authors as described in the Methods. Analysis code will be deposited in a public repository upon publication.

## Acknowledgments

This work was supported by the Swiss National Science Foundation (SNSF), grant ID: 315230-219587. The SNSF also covered the open access publication costs.

## Appendix

### Supplementary Tables

**Table 1:** For six of 34 traits (17.6%) the cross-sex genetic correlation is nominally significantly different from 1 indicative of sex-different genetic architecture in the respective trait. After FDR correction, the cross-sex genetic correlation of testosterone and WHR remain significantly different from 1.

| Trait | cross-sex rG (95% CI) | *p*-value |
| --- | --- | --- |
| BMI | 0.91 (0.86, 0.97) | 5.41E-02 |
| GGT | 0.88 (0.78, 0.98) | 9.24E-02 |
| IGF1 | 0.88 (0.78, 0.99) | 1.57E-01 |
| LDL | 0.82 (0.65, 1) | 2.00E-01 |
| SHBG | 0.84 (0.71, 0.96) | 9.12E-02 |
| SMK never | 0.91 (0.83, 0.98) | 9.24E-02 |
| Testosterone | 0 (-0.08, 0.07) | 3.16E-152 |
| WHR | 0.73 (0.67, 0.78) | 7.89E-23 |

**Supplementary Table 1:** Overview of Trait names, abbreviations, number of combined, male and female individuals and Field ID used to extract phenotypes from the UK Biobank. ^1)^For diseases COPD and GERD, doctor diagnosed individuals without cancer were selected. Individuals listed under Field IDs 20002-1112 (COPD self-reported) and 20002-1159 (GERD self-reported) were removed. ^2)^WHR was calculated by dividing waist circumference (Field ID 48) by hip circumference (Field ID 49). A GWAS was conducted to obtain summary statistics.

| Abbreviation | Trait name | Number of individuals |  |  | Field ID UK Biobank | Trait type |
| --- | --- | --- | --- | --- | --- | --- |
|  |  | Combined | Women | Men |  |  |
| Alcohol | Alcohol | 146072 | 79123 | 66949 | 100022 | continuous |
| ALP | Alkaline Phosphatase | 322617 | 173158 | 149459 | 30610 | continuous |
| ApoB | Apolipoprotein B | 321071 | 172561 | 148510 | 30640 | continuous |
| Asthma | Asthma | 267450 | 142195 | 125246 | 20002-1111 | binary |
| Basophil | Basophil count | 327694 | 175664 | 152030 | 30160 | continuous |
| BMI | Body mass index | 336378 | 180681 | 155697 | 21001 | continuous |
| Cholesterol | Cholesterol | 322605 | 173141 | 149464 | 30690 | continuous |
| COPD | Chronic Obstructive Pulmonary Disease | 290753 | 154848 | 135896 | 41270-J44<sup>1)</sup> | binary |
| CRP | C-reactive protein | 321928 | 172838 | 149090 | 30710 | continuous |
| CysC | Cystatin C | 322582 | 173133 | 149449 | 30720 | continuous |
| Edu | Age when finish education | 230065 | 125925 | 104140 | 845 | continuous |
| Eosinophil | Eosinophil | 327694 | 175664 | 152030 | 30150 | continuous |
| FEV1 | Forced expiratory volume in 1 second | 309854 | 167623 | 142231 | 20153 | continuous |
| FVC | Forced vital capacity | 309854 | 167623 | 142231 | 20151 | continuous |
| GERD | Gastroesophageal reflux disease | 283329 | 151192 | 132128 | 41270-K21.0, K21.9<sup>1)</sup> | binary |
| GGT | Gamma glutamyl transferase | 322446 | 173074 | 149372 | 30730 | continuous |
| Glucose | Glucose | 295955 | 157688 | 138267 | 30740 | continuous |
| Hb | Haemoglobin | 328246 | 175953 | 152293 | 30020 | continuous |
| HbA1c | Glycated haemoglobin | 322585 | 173217 | 149368 | 30750 | continuous |
| HDL | High-density lipoprotein cholesterol | 296132 | 157781 | 138351 | 30760 | continuous |
| Height | Standing height | 336725 | 180858 | 155867 | 50 | continuous |
| IGF1 | Insulin-like growth factor 1 | 320941 | 172234 | 148707 | 30770 | continuous |
| LDL | Low-density lipoprotein cholesterol | 322029 | 172872 | 149157 | 30780 | continuous |
| Neutrophil | Neutrophil | 327694 | 175664 | 152030 | 30140 | continuous |
| Oestradiol | Oestradiol | 51218 | 38162 | 13056 | 30800 | continuous |
| SHBG | Sex-hormone binding globulin | 293464 | 156090 | 137374 | 30830 | continuous |
| SMK current | Smoking status: current | 337423 | 181203 | 156220 | 20116-2 | binary |
| SMK never | Smoking status: never | 337423 | 181203 | 156220 | 20116-0 | binary |
| SMK former | Smoking status: former | 337423 | 181203 | 156220 | 20116-1 | binary |
| Test | Testosterone | 292994 | 144933 | 148061 | 30850 | continuous |
| TG | Triglycerides | 322343 | 173035 | 149308 | 30870 | continuous |
| Urate | Urate | 322196 | 172916 | 149280 | 30880 | continuous |
| VitD | Vitamin D | 308703 | 163522 | 145181 | 30890 | continuous |
| WHR | Waist-hip ratio | 336811 | 180871 | 155940 | 48/49<sup>2)</sup> | continuous |

**Supplementary Table 2:** Sex-stratified mean and standard deviation (SD) of the 34 investigated traits. g: gram; U: units of measurement; L: liter; umol: micromole.

| Trait name | Women |  | Men |  | Unit |
| --- | --- | --- | --- | --- | --- |
|  | Mean | SD | Mean | SD |  |
| Alcohol | 12.2 | 16.05 | 22.11 | 25.33 | g |
| Alkaline Phosphatase | 85.06 | 27.66 | 82.02 | 24.81 | U/L |
| Apolipoprotein B | 1.04 | 0.24 | 1.03 | 0.24 | g/L |
| Asthma | 1.1 | 0.3 | 1.08 | 0.27 | 10<sup>9</sup> cells/L |
| Basophil count | 0.04 | 0.05 | 0.03 | 0.05 | 10<sup>9</sup> cells/L |
| Body mass index | 27.03 | 5.13 | 27.83 | 4.22 | kg/m<sup>2</sup> |
| Cholesterol | 5.9 | 1.12 | 5.5 | 1.12 | mmol/L |
| Chronic Obstructive Pulmonary Disease | 1.03 | 0.16 | 1.04 | 0.19 | - |
| C-reactive protein | 2.69 | 4.33 | 2.46 | 4.37 | mg/L |
| Cystatin C | 0.88 | 0.16 | 0.94 | 0.18 | mg/L |
| Age when finish education | 16.44 | 2.72 | 16.46 | 3.12 | years |
| Eosinophil | 0.16 | 0.13 | 0.19 | 0.14 | 10<sup>9</sup> cells/L |
| Forced expiratory volume in 1 second | 2.33 | 0.53 | 3.22 | 0.74 | L |
| Forced vital capacity | 3.11 | 0.67 | 4.35 | 0.92 | L |
| Gastroesophageal reflux disease | 1.09 | 0.28 | 1.08 | 0.27 | - |
| Gamma glutamyl transferase | 30.32 | 34.06 | 45.47 | 47.85 | U/L |
| Glucose | 5.07 | 1.04 | 5.18 | 1.36 | mmol/L |
| Haemoglobin | 13.53 | 0.95 | 15.01 | 1.01 | g/dL |
| Glycated haemoglobin | 35.67 | 5.7 | 36.31 | 7.31 | mmol/L |
| High-density lipoprotein cholesterol | 1.6 | 0.38 | 1.29 | 0.31 | mmol/L |
| Standing height | 162.68 | 6.21 | 175.93 | 6.76 | cm |
| Insulin-like growth factor 1 | 20.94 | 5.72 | 21.93 | 5.52 | nmol/L |
| Low-density lipoprotein cholesterol | 3.64 | 0.87 | 3.49 | 0.86 | mmol/L |
| Neutrophil | 4.21 | 1.38 | 4.29 | 1.43 | 10<sup>9</sup> cells/L |
| Oestradiol | 537.33 | 475 | 222.94 | 79.17 | pmol/L |
| Sex-hormone binding globulin | 62.29 | 30.85 | 40.04 | 16.8 | mmol/L |
| Smoking status: current | 0.08 | 0.28 | 0.11 | 0.32 | - |
| Smoking status: never | 0.59 | 0.49 | 0.49 | 0.5 | - |
| Smoking status: former | 0.32 | 0.46 | 0.39 | 0.49 | - |
| Testosterone | 1.12 | 0.63 | 11.97 | 3.7 | mmol/L |
| Triglycerides | 1.56 | 0.85 | 1.98 | 1.14 | mmol/L |
| Urate | 270.99 | 66.02 | 354.41 | 71.37 | umol/L |
| Vitamin D | 49.76 | 20.86 | 49.8 | 20.96 | nmol/L |
| Waist-hip ratio | 0.82 | 0.07 | 0.94 | 0.06 | - |

**Supplementary Table 3:**
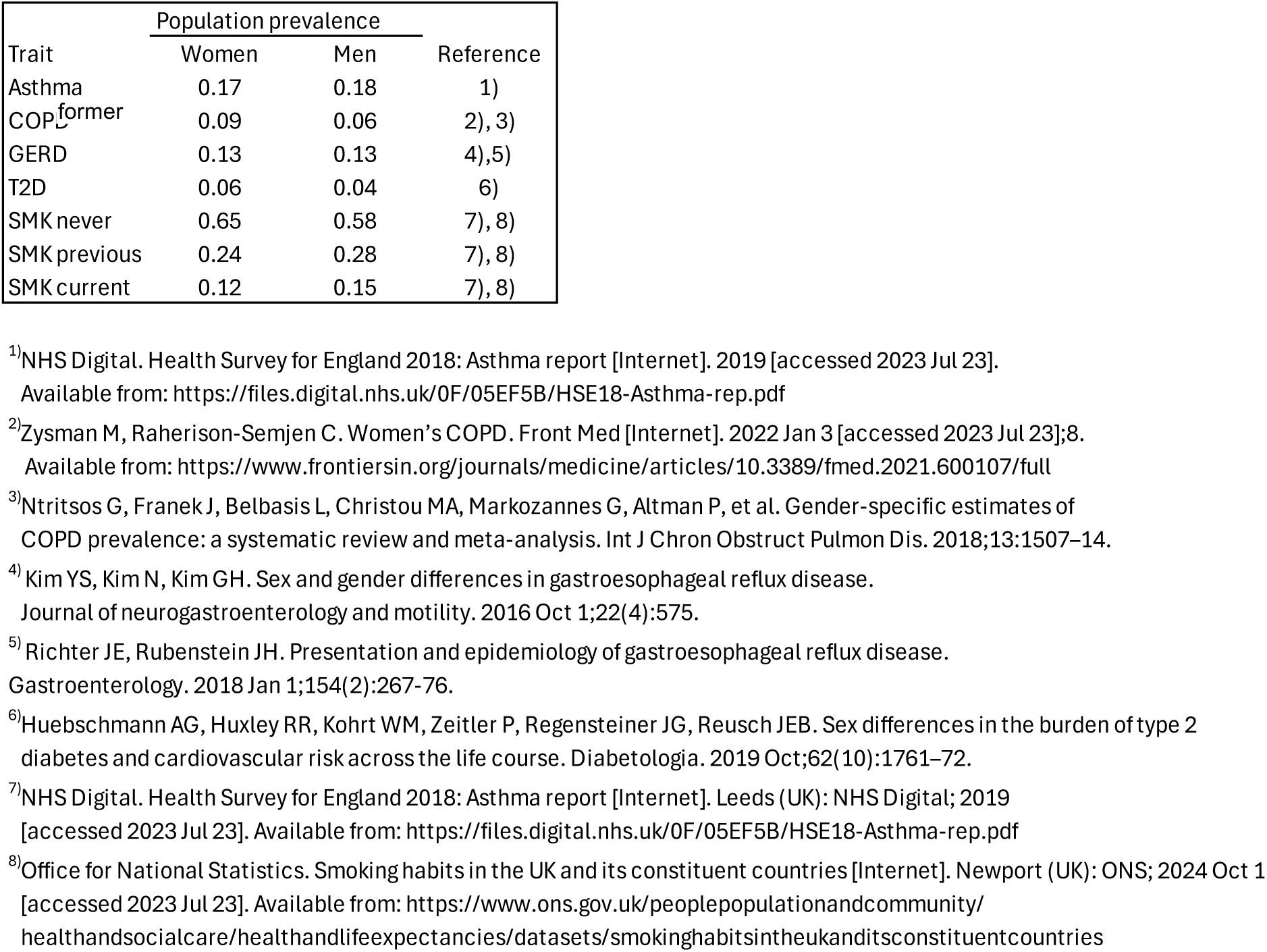
Population prevalences used in LDSC for SNP-heritability estimation from the observed scale to the liability scale for the binary traits asthma, COPD, GERD, T2D and smoking behaviour.

**Supplementary Table 4:**
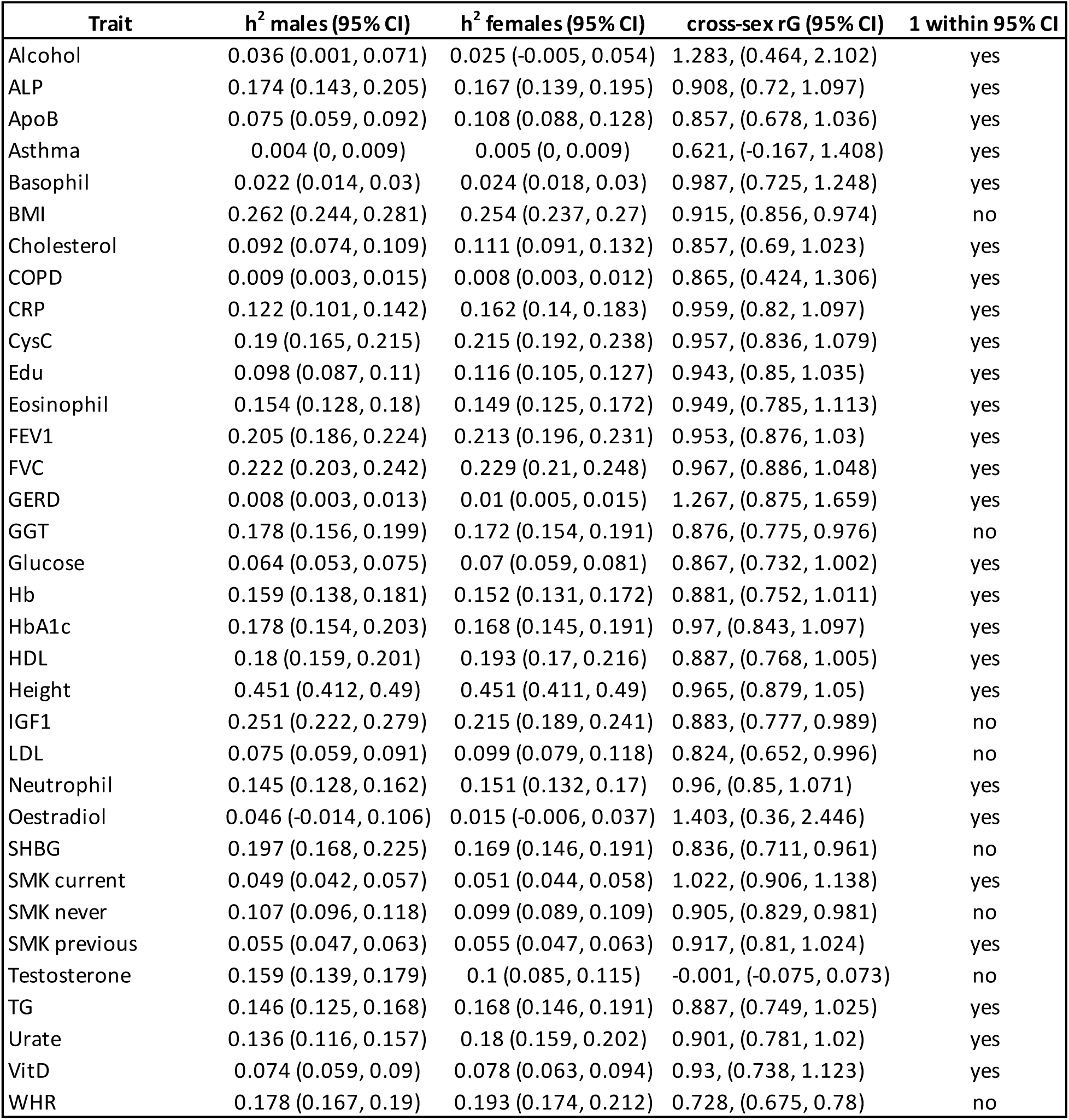
Sex-stratified narrow-sense heritability (h^2^ males, h^2^ females) and genetic correlation between males and females within one trait (intra-trait rG) with 95% confidence interval (95% CI); Last column indicates if intra-trait genetic correlation between males and females is significantly different from 1, which gives insight if males and females share the same genetic mechanisms affecting the respective trait; yes: 1 is not within the 95% CI of intra-trait rG, no: 1 is not within the 95%.

**Supplementary Table 5:**
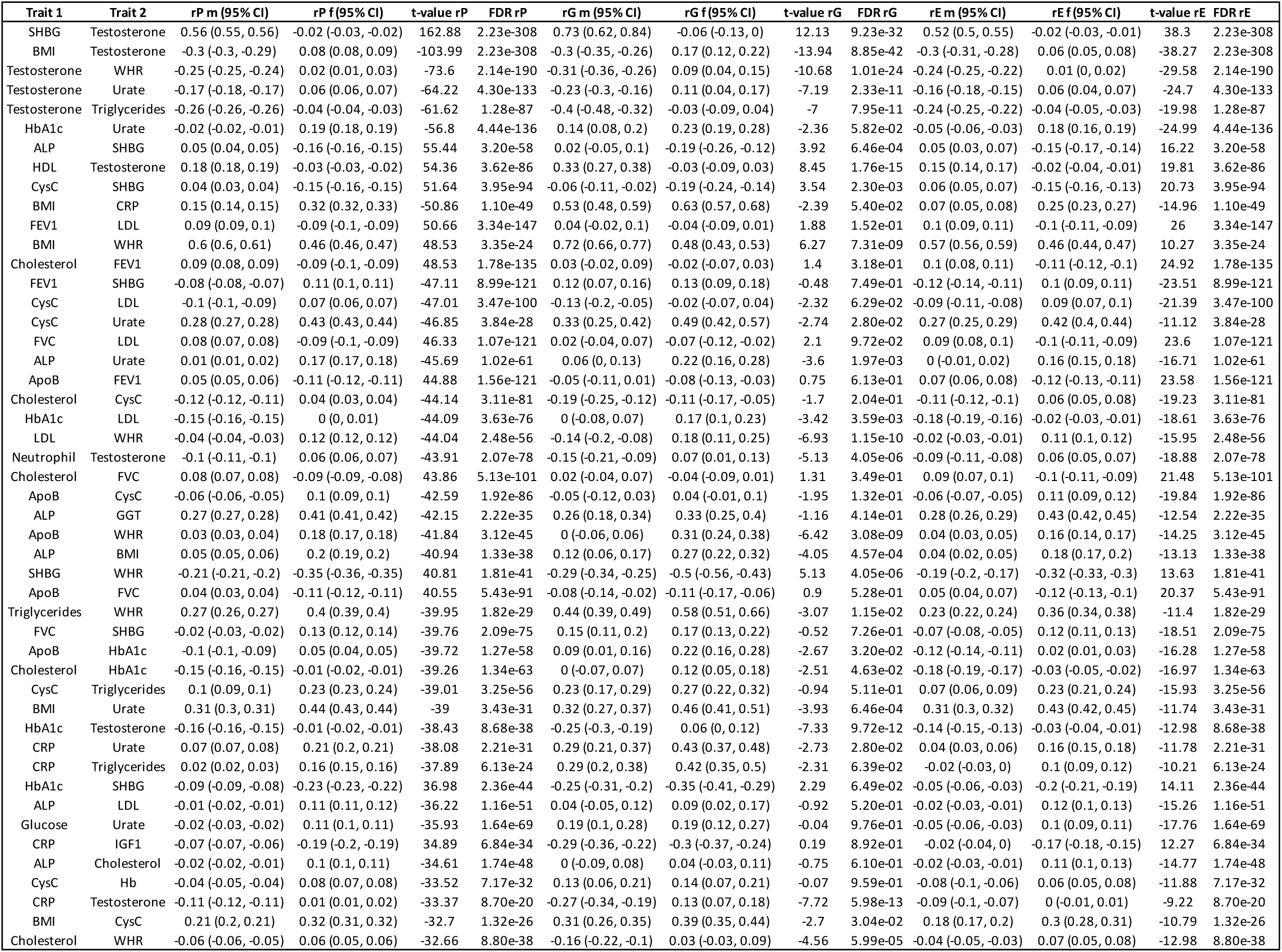
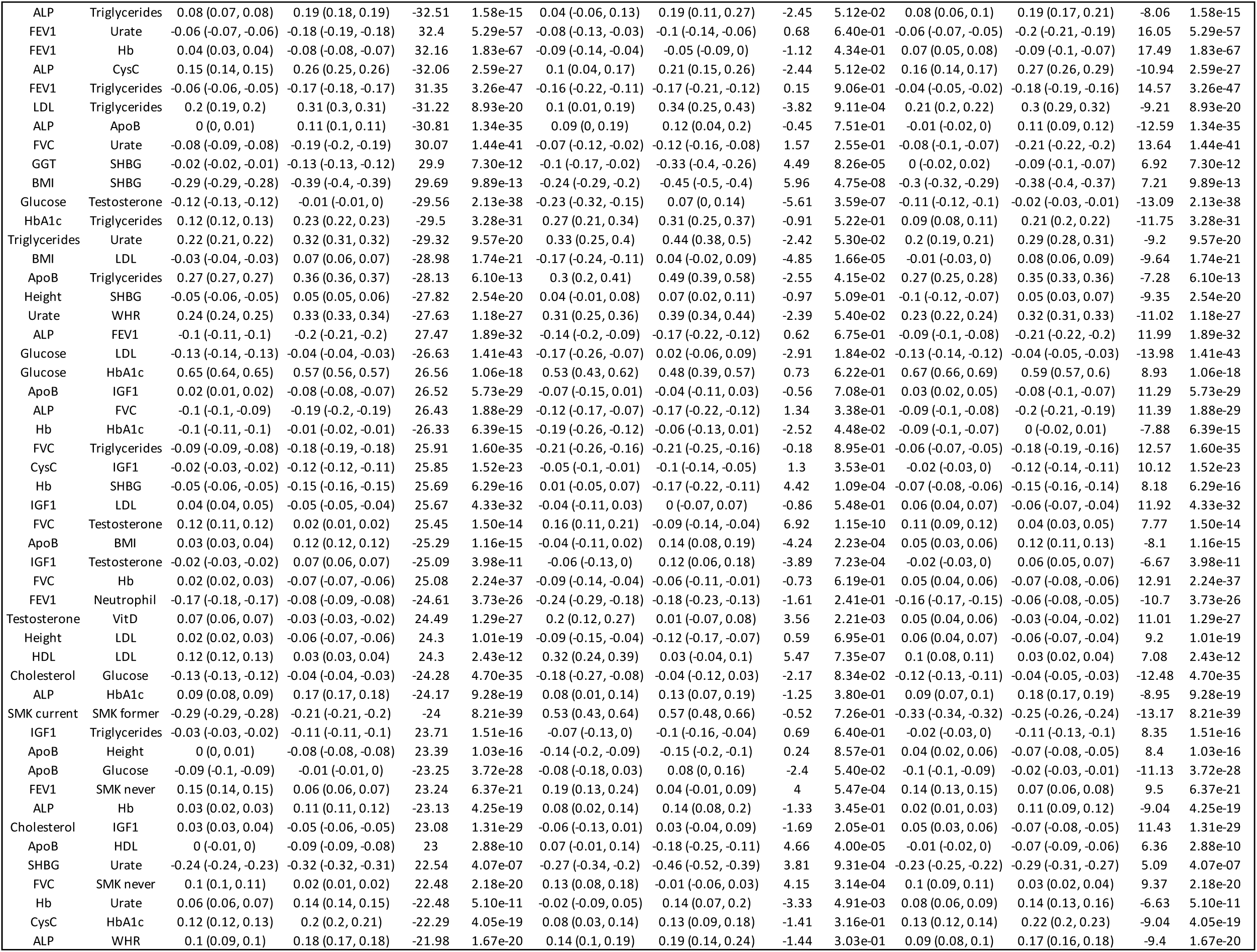
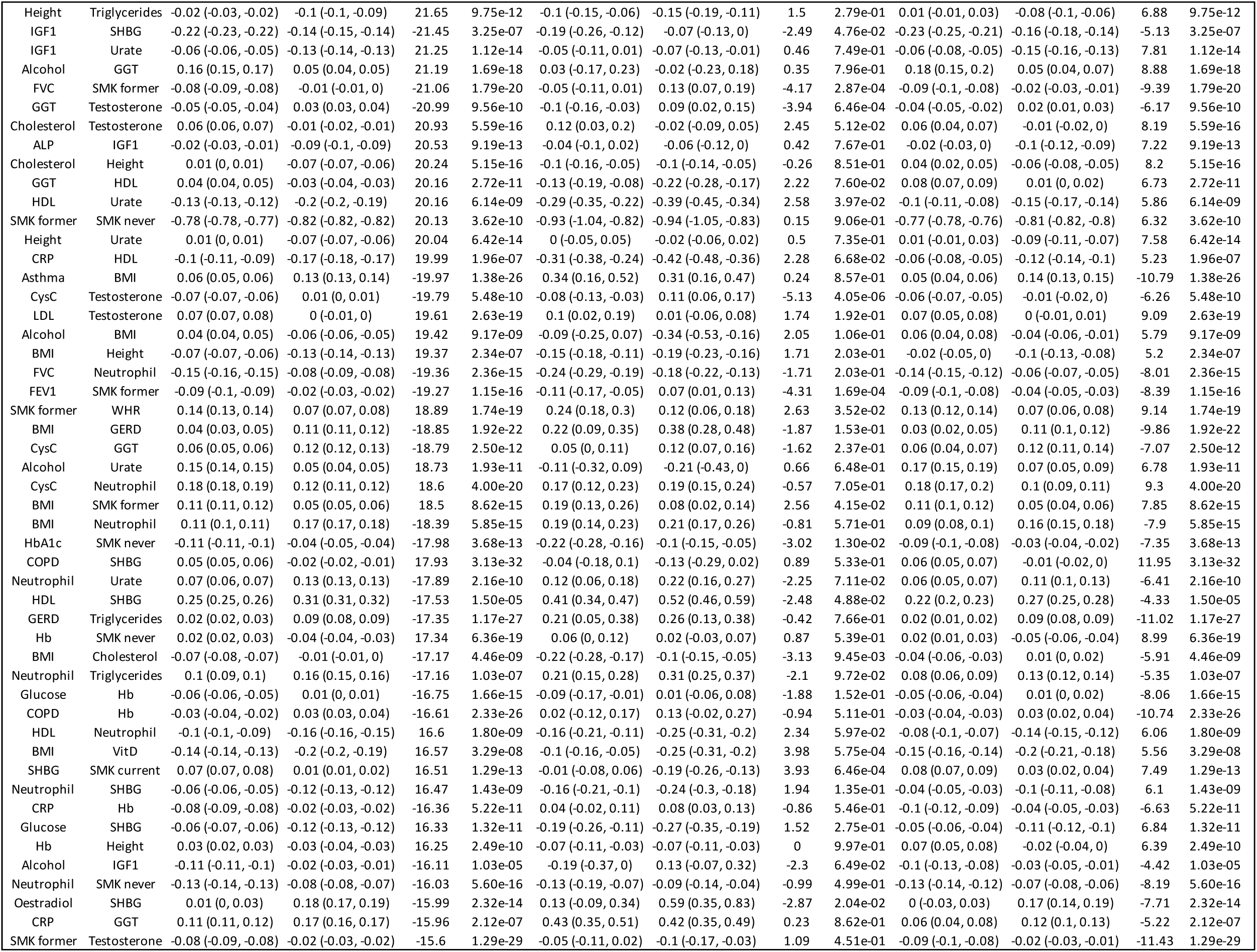
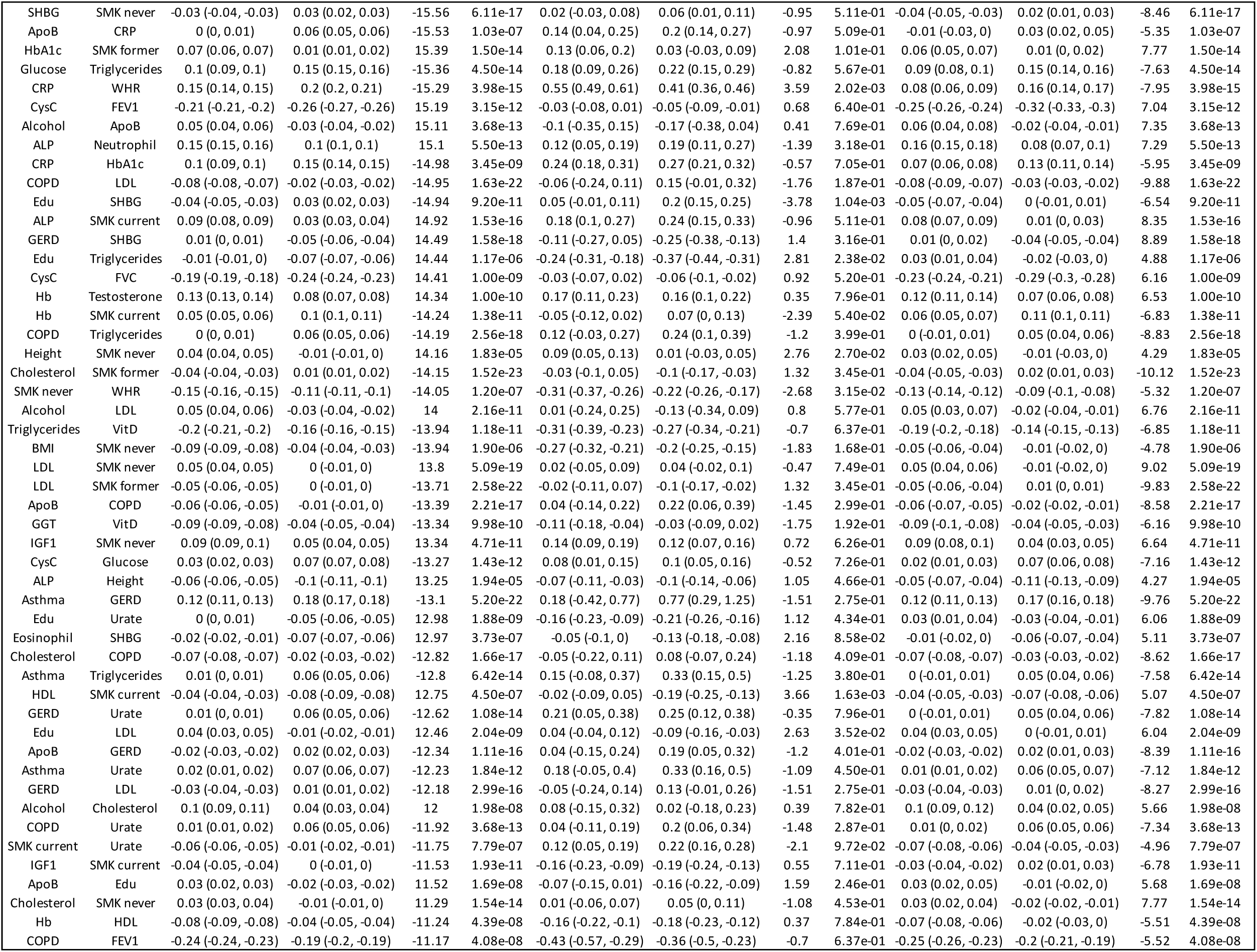
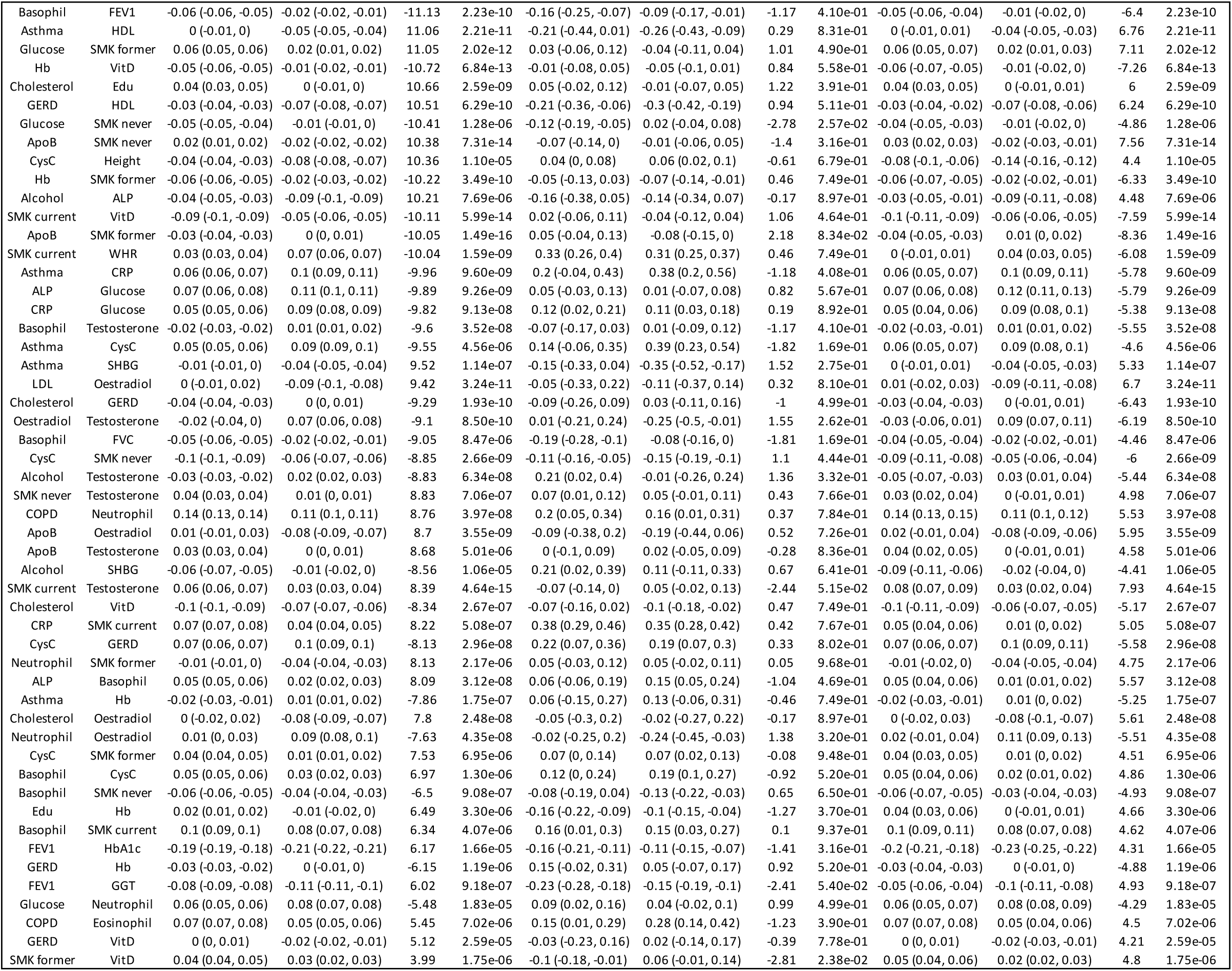
Phenotypic correlation of males (rP m) and females (rP f) with 95% confidence interval (95% CI) *t*-value (*t*-value rP) and corrected *p*-value (FDR rP) to evaluate whether there is a significant difference between male and female phenotypic correlation, genetic correlation of males (rG m) and females (rG f) with 95% confidence interval (95% CI) *t*-value (*t*-value rG) and corrected *p*-value (FDR rG) to evaluate whether there is a significant difference between male and female genetic correlation; environmental correlation of males (rE m) and emales (rE f) with 95% confidence interval (95% CI) *t*-value (*t*-value rE) and corrected *p*-value (FDR rE) to evaluate whether there is a significant difference between male and female environmental correlation;

**Supplementary Figure 1:**
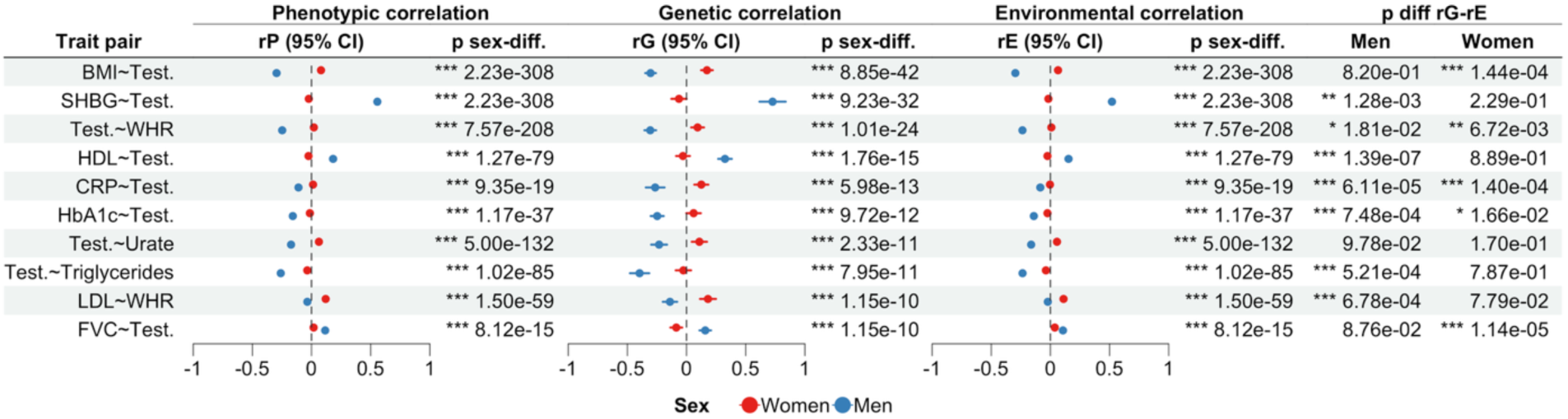
Showing 10 trait pairs with the **strongest sex-different rG**. Columns show phenotypic correlation (rP), genetic correlation (rG), and environmental correlation (rE) with 95% confidence intervals (95% CI), their respective *p*-value of the *t*-test between the correlation between men and women (p sex-diff.), and *p*-value of the *t*-test between the rG and rE (p diff rG-rE), separate for men and women.

**Supplementary Figure 2:**
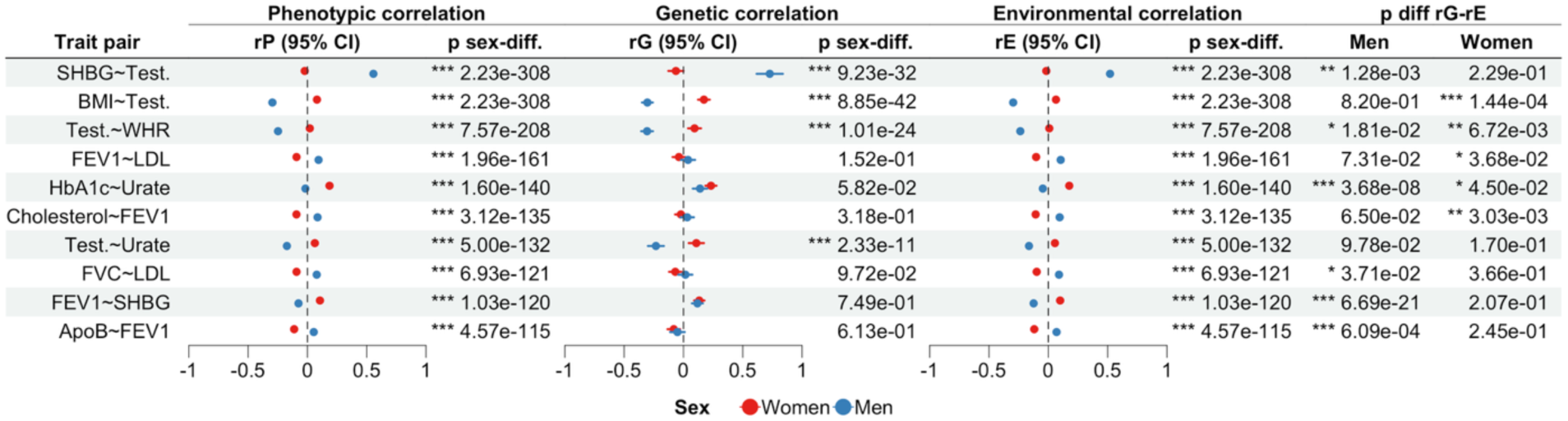
Showing 10 trait pairs with the strongest **sex-different rE.** Columns show phenotypic correlation (rP), genetic correlation (rG), and environmental correlation (rE) with 95% confidence intervals (95% CI), *p*-value (*t*-test) of sex-difference in the respective correlation (p sex-diff.) determined by *t*-test, and *p*-value of difference between rG and rE (p diff rG-rE) determined by *t*-test in men and women.

**Supplementary Figure 3:**
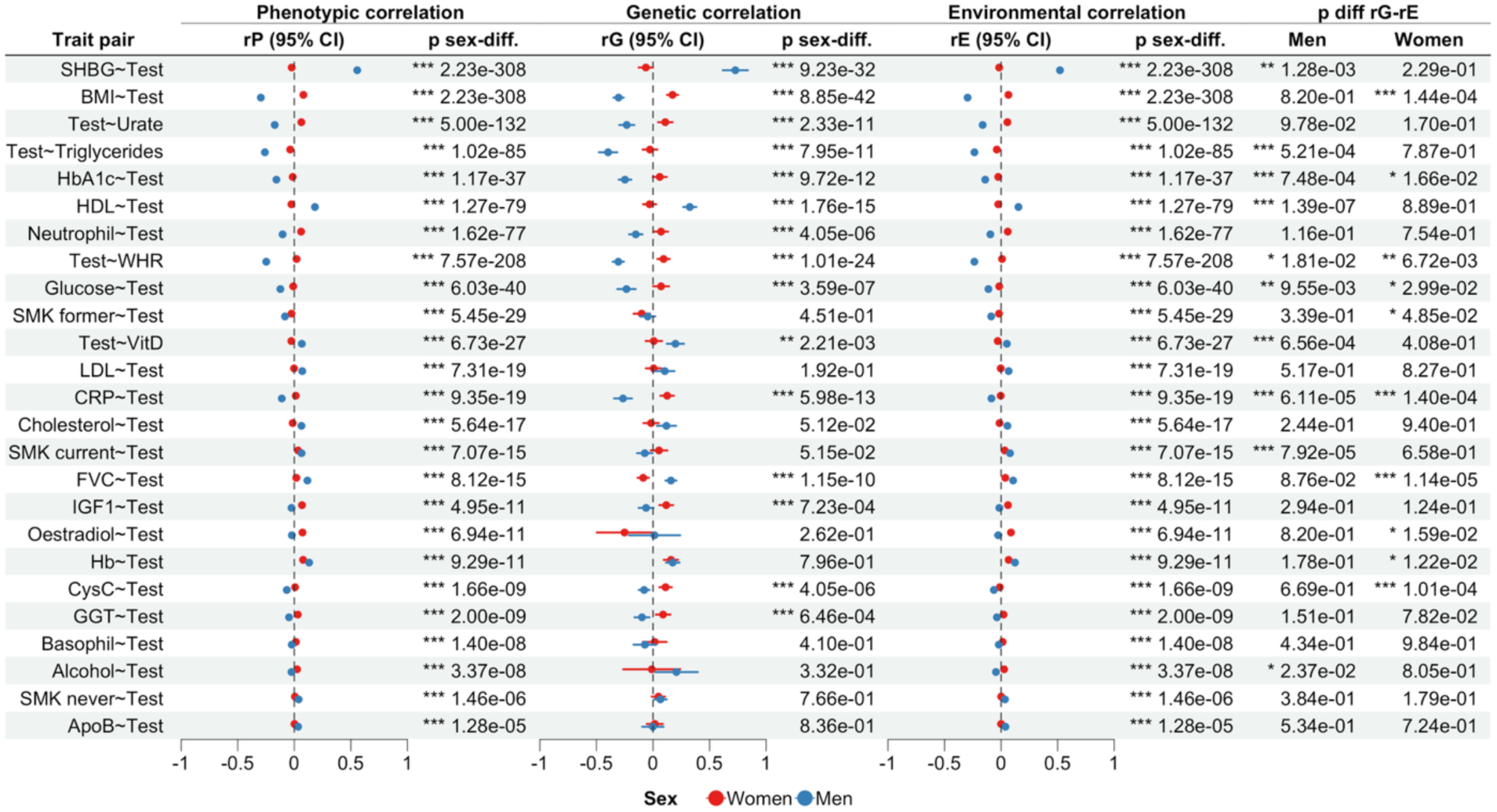
Showing traits associated with testosterone. Columns show phenotypic correlation (rP), genetic correlation (rG), and environmental correlation (rE) with 95% confidence intervals (95% CI), *p*-value (*t*-test) of sex-difference in the respective correlation (p sex-diff.) determined by *t*-test, and *p*-value of difference between rG and rE (p diff rG-rE) determined by *t*-test in men and women.

**Supplementary Table 6:**
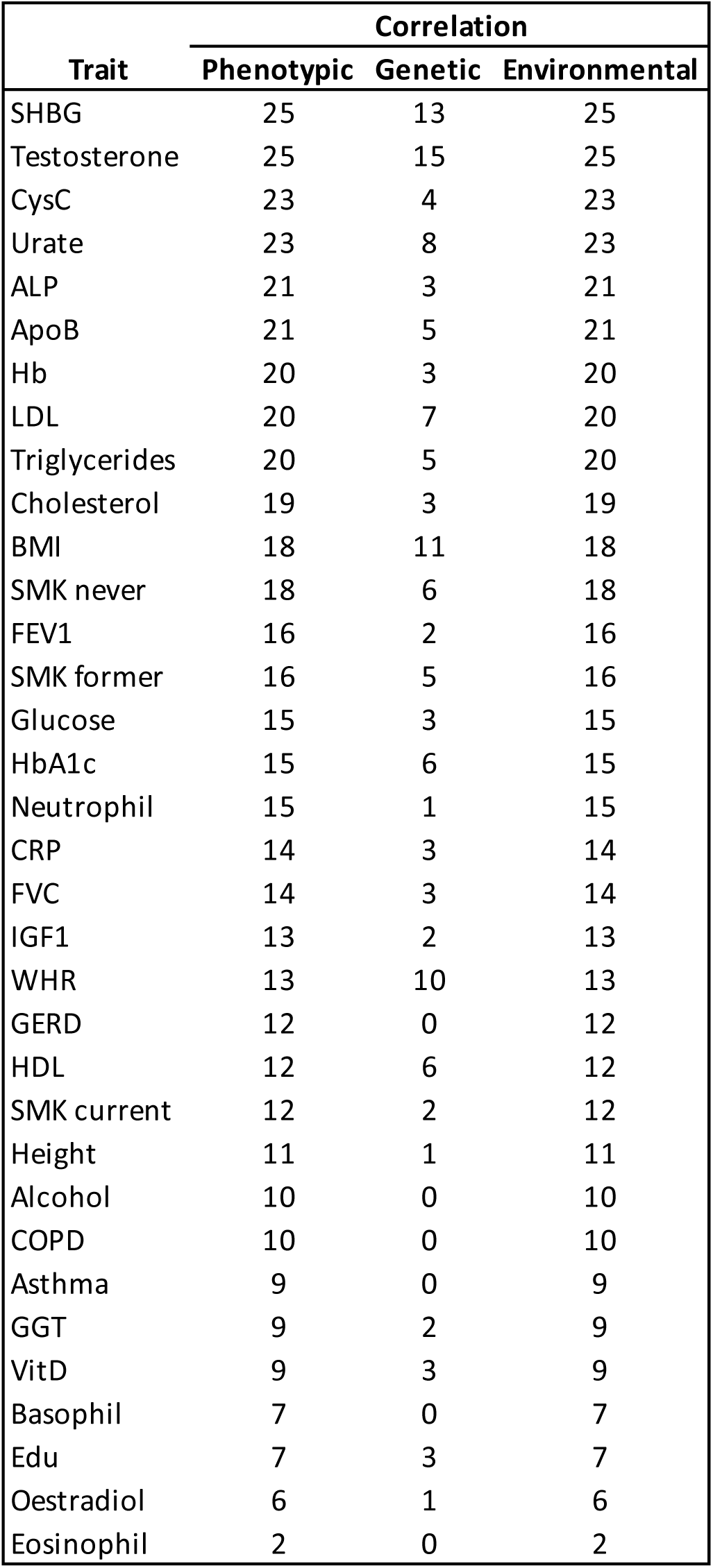
Frequency of traits being part of pairs showing significant sex-different phenotypic (left), genetic (middle) and environmental correlation.

## References

1. Ober C, Loisel DA, Gilad Y. Sex-specific genetic architecture of human disease. Nat Rev Genet. 2008 Dec;9(12):911–22.

2. Randall JC, Winkler TW, Kutalik Z, Berndt SI, Jackson AU, Monda KL, et al. Sex-stratified genome-wide association studies including 270,000 individuals show sexual dimorphism in genetic loci for anthropometric traits. PLoS Genet. 2013 Jun;9(6):e1003500.

3. Ngo ST, Steyn FJ, McCombe PA. Gender differences in autoimmune disease. Front Neuroendocrinol. 2014 Aug;35(3):347–69.

4. Voskuhl R. Sex differences in autoimmune diseases. Biol Sex Differ. 2011 Jan 4;2(1):1.

5. McLean CP, Asnaani A, Litz BT, Hofmann SG. Gender differences in anxiety disorders: prevalence, course of illness, comorbidity and burden of illness. J Psychiatr Res. 2011 Aug;45(8):1027–35.

6. Salk RH, Hyde JS, Abramson LY. Gender differences in depression in representative national samples: Meta-analyses of diagnoses and symptoms. Psychol Bull. 2017 Aug;143(8):783–822.

7. Weidner G. Why do men get more heart disease than women? An international perspective. J Am Coll Health J ACH. 2000 May;48(6):291–4.

8. Kautzky-Willer A, Leutner M, Harreiter J. Sex differences in type 2 diabetes. Diabetologia. 2023;66(6):986–1002.

9. Fuseini H, Newcomb DC. Mechanisms Driving Gender Differences in Asthma. Curr Allergy Asthma Rep. 2017 Mar;17(3):19.

10. Kynyk JA, Mastronarde JG, McCallister JW. Asthma, the sex difference. Curr Opin Pulm Med. 2011 Jan;17(1):6–11.

11. Walli-Attaei M, Rosengren A, Rangarajan S, Breet Y, Abdul-Razak S, Sharief WA, et al. Metabolic, behavioural, and psychosocial risk factors and cardiovascular disease in women compared with men in 21 high-income, middle-income, and low-income countries: an analysis of the PURE study. The Lancet. 2022 Sep;400(10355):811–21.

12. Rawlik K, Canela-Xandri O, Tenesa A. Evidence for sex-specific genetic architectures across a spectrum of human complex traits. Genome Biol. 2016 Jul 29;17(1):166.

13. Weiss LA, Pan L, Abney M, Ober C. The sex-specific genetic architecture of quantitative traits in humans. Nat Genet. 2006 Feb;38(2):218–22.

14. Porcu E, Claringbould A, Weihs A, Lepik K, BIOS Consortium, Richardson TG, et al. Limited evidence for blood eQTLs in human sexual dimorphism. Genome Med. 2022 Aug 11;14(1):89.

15. Patsopoulos NA, Tatsioni A, Ioannidis JPA. Claims of sex differences: an empirical assessment in genetic associations. JAMA. 2007 Aug 22;298(8):880–93.

16. Elgart M, Goodman MO, Isasi C, Chen H, Morrison AC, de Vries PS, et al. Correlations between complex human phenotypes vary by genetic background, gender, and environment. Cell Rep Med. 2022 Dec 12;3(12):100844.

17. Baye TM, Abebe T, Wilke RA. Genotype–environment interactions and their translational implications. Pers Med. 2011 Jan;8(1):59–70.

18. Bycroft C, Freeman C, Petkova D, Band G, Elliott LT, Sharp K, et al. The UK Biobank resource with deep phenotyping and genomic data. Nature. 2018 Oct;562(7726):203–9.

19. Benesty J, Chen J, Huang Y, Cohen I. Pearson Correlation Coefficient. In: Cohen I, Huang Y, Chen J, Benesty J, editors. Noise Reduction in Speech Processing [Internet]. Berlin, Heidelberg: Springer; 2009 [cited 2023 Sep 13]. p. 1–4. (Springer Topics in Signal Processing). Available from: 10.1007/978-3-642-00296-0_5

20. Maurage P, Heeren A, Pesenti M. Does chocolate consumption really boost Nobel Award chances? The peril of over-interpreting correlations in health studies. J Nutr. 2013 Jun;143(6):931–3.

21. Kim TK. T test as a parametric statistic. Korean J Anesthesiol. 2015 Nov 25;68(6):540–6.

22. Bulik-Sullivan BK, Loh PR, Finucane H, Ripke S, Yang J, Patterson N, et al. LD Score Regression Distinguishes Confounding from Polygenicity in Genome-Wide Association Studies. Nat Genet. 2015 Mar;47(3):291–5.

23. Grotzinger AD, Rhemtulla M, de Vlaming R, Ritchie SJ, Mallard TT, Hill WD, et al. Genomic structural equation modelling provides insights into the multivariate genetic architecture of complex traits. Nat Hum Behav. 2019 May;3(5):513–25.

24. Neale lab [Internet]. [cited 2023 Dec 12]. UK Biobank. Available from: http://www.nealelab.is/uk-biobank

25. Sodini SM, Kemper KE, Wray NR, Trzaskowski M. Comparison of Genotypic and Phenotypic Correlations: Cheverud’s Conjecture in Humans. Genetics. 2018 Jul 1;209(3):941–8.

26. Tang M, Wang T, Zhang X. A review of SNP heritability estimation methods. Brief Bioinform. 2022 May 1;23(3):bbac067.

27. Concepts, estimation and interpretation of SNP-based heritability | Nature Genetics [Internet]. [cited 2024 Oct 25]. Available from: https://www.nature.com/articles/ng.3941

28. Bernabeu E, Canela-Xandri O, Rawlik K, Talenti A, Prendergast J, Tenesa A. Sex differences in genetic architecture in the UK Biobank. Nat Genet. 2021 Sep;53(9):1283–9.

29. Ni G, Moser G, Schizophrenia Working Group of the Psychiatric Genomics Consortium, Wray NR, Lee SH. Estimation of Genetic Correlation via Linkage Disequilibrium Score Regression and Genomic Restricted Maximum Likelihood. Am J Hum Genet. 2018 Jun 7;102(6):1185–94.

30. Ruth KS, Day FR, Tyrrell J, Thompson DJ, Wood AR, Mahajan A, et al. Using human genetics to understand the disease impacts of testosterone in men and women. Nat Med. 2020 Feb;26(2):252–8.

31. Pulit SL, Stoneman C, Morris AP, Wood AR, Glastonbury CA, Tyrrell J, et al. Meta-analysis of genome-wide association studies for body fat distribution in 694 649 individuals of European ancestry. Hum Mol Genet. 2019 Jan 1;28(1):166–74.

32. Sanderson E, Glymour MM, Holmes MV, Kang H, Morrison J, Munafò MR, et al. Mendelian randomization. Nat Rev Methods Primer. 2022 Feb 10;2:6.

33. Smith GD, Ebrahim S. “Mendelian randomization”: can genetic epidemiology contribute to understanding environmental determinants of disease? Int J Epidemiol. 2003 Feb;32(1):1–22.

34. Handelsman DJ, Hirschberg AL, Bermon S. Circulating Testosterone as the Hormonal Basis of Sex Differences in Athletic Performance. Endocr Rev. 2018 Jul 13;39(5):803–29.

35. Lundberg S. Educational Gender Gaps. South Econ J. 2020 Oct;87(2):416–39.

36. Eveleth PB, Tanner JM. Worldwide Variation in Human Growth [Internet]. Cambridge: Cambridge University Press; 1991 [cited 2024 Oct 15]. Available from: https://www.cambridge.org/core/books/worldwide-variation-in-human-growth/0E6E8B888E2F2B9F4F17112935BA1928

37. Winters SJ, Gogineni J, Karegar M, Scoggins C, Wunderlich CA, Baumgartner R, et al. Sex Hormone-Binding Globulin Gene Expression and Insulin Resistance. J Clin Endocrinol Metab. 2014 Dec 1;99(12):E2780–8.

38. Shungin D, Winkler TW, Croteau-Chonka DC, Ferreira T, Locke AE, Mägi R, et al. New genetic loci link adipose and insulin biology to body fat distribution. Nature. 2015 Feb 12;518(7538):187–96.

39. Flynn E, Tanigawa Y, Rodriguez F, Altman RB, Sinnott-Armstrong N, Rivas MA. Sex-specific genetic effects across biomarkers. Eur J Hum Genet. 2021 Jan;29(1):154– 63.

40. Huang Y, Shan Y, Zhang W, Lee AM, Li F, Stranger BE, et al. Deciphering genetic causes for sex differences in human health through drug metabolism and transporter genes. Nat Commun. 2023 Jan 12;14(1):175.

41. Sterling T, Weinkam J. The confounding of occupation and smoking and its consequences. Soc Sci Med. 1990 Jan 1;30(4):457–67.

42. Abdellaoui A, Dolan CV, Verweij KJH, Nivard MG. Gene–environment correlations across geographic regions affect genome-wide association studies. Nat Genet. 2022 Sep;54(9):1345–54.

43. Young AS. Genome-wide association studies have problems due to confounding: Are family-based designs the answer? PLOS Biol. 2024 Apr 12;22(4):e3002568.

44. Nivard MG, Belsky DW, Harden KP, Baier T, Andreassen OA, Ystrøm E, et al. More than nature and nurture, indirect genetic effects on children’s academic achievement are consequences of dynastic social processes. Nat Hum Behav. 2024 Apr;8(4):771–8.

45. Krakowsky Y, Grober ED. Testosterone Deficiency - Establishing A Biochemical Diagnosis. EJIFCC. 2015 Mar;26(2):105.

46. Leinonen JT, Mars N, Lehtonen LE, Ahola-Olli A, Ruotsalainen S, Lehtimäki T, et al. Genetic analyses implicate complex links between adult testosterone levels and health and disease. Commun Med. 2023 Jan 18;3(1):1–15.

47. Keenan RT. The biology of urate. Semin Arthritis Rheum. 2020 Jun;50(3S):S2–10.

48. Wang Y, Charchar FJ. Establishment of sex difference in circulating uric acid is associated with higher testosterone and lower sex hormone-binding globulin in adolescent boys. Sci Rep. 2021 Aug 30;11(1):17323.

49. Liu Y, Elsworth B, Erola P, Haberland V, Hemani G, Lyon M, et al. EpiGraphDB: a database and data mining platform for health data science. Bioinforma Oxf Engl. 2021 Jun 9;37(9):1304–11.

50. Rasheed H, Hsu A, Dalbeth N, Stamp LK, McCormick S, Merriman TR. The relationship of apolipoprotein B and very low density lipoprotein triglyceride with hyperuricemia and gout. Arthritis Res Ther. 2014 Nov 29;16(6):495.

51. Levinson T, Wasserman A. C-Reactive Protein Velocity (CRPv) as a New Biomarker for the Early Detection of Acute Infection/Inflammation. Int J Mol Sci. 2022 Jan;23(15):8100.

52. Nari F, Jang BN, Kim GR, Park EC, Jang SI. Synergistic Effects and Sex Differences in Anthropometric Measures of Obesity and Elevated High-Sensitivity C-Reactive Protein Levels. Int J Environ Res Public Health. 2020 Nov;17(21):8279.

53. Moreno-Llamas A, García-Mayor J, De la Cruz-Sánchez E. Gender inequality is associated with gender differences and women participation in physical activity. J Public Health Oxf Engl. 2022 Dec 1;44(4):e519–26.

54. Meier HE, Konjer MV, Krieger J. Women in International Elite Athletics: Gender (in)equality and National Participation. Front Sports Act Living. 2021;3:709640.

55. Eyth E, Naik R. Hemoglobin A1C. In: StatPearls [Internet]. Treasure Island (FL): StatPearls Publishing; 2024 [cited 2024 Mar 21]. Available from: http://www.ncbi.nlm.nih.gov/books/NBK549816/

56. Glycated haemoglobin (HbA1c) for the diagnosis of diabetes. In: Use of Glycated Haemoglobin (HbA1c) in the Diagnosis of Diabetes Mellitus: Abbreviated Report of a WHO Consultation [Internet]. World Health Organization; 2011 [cited 2024 Mar 21]. Available from: https://www.ncbi.nlm.nih.gov/books/NBK304271/

57. Luo J, Yang H, Song BL. Mechanisms and regulation of cholesterol homeostasis. Nat Rev Mol Cell Biol. 2020 Apr;21(4):225–45.

58. Ference BA, Yoo W, Alesh I, Mahajan N, Mirowska KK, Mewada A, et al. Effect of Long-Term Exposure to Lower Low-Density Lipoprotein Cholesterol Beginning Early in Life on the Risk of Coronary Heart Disease: A Mendelian Randomization Analysis. J Am Coll Cardiol. 2012 Dec 25;60(25):2631–9.

59. Gencer B, Marston NA, Im K, Cannon CP, Sever P, Keech A, et al. Efficacy and safety of lowering LDL cholesterol in older patients: a systematic review and meta-analysis of randomised controlled trials. Lancet Lond Engl. 2020 Nov 21;396(10263):1637–43.

60. Freeman DJ, Norrie J, Sattar N, Neely RD, Cobbe SM, Ford I, et al. Pravastatin and the development of diabetes mellitus: evidence for a protective treatment effect in the West of Scotland Coronary Prevention Study. Circulation. 2001 Jan 23;103(3):357–62.

61. Goldenberg N, Glueck C. Efficacy, effectiveness and real life goal attainment of statins in managing cardiovascular risk. Vasc Health Risk Manag. 2009;5:369–76.

62. Feingold KR. Cholesterol Lowering Drugs. In: Feingold KR, Ahmed SF, Anawalt B, Blackman MR, Boyce A, Chrousos G, et al., editors. Endotext [Internet]. South Dartmouth (MA): MDText.com, Inc.; 2000 [cited 2025 Sep 23]. Available from: http://www.ncbi.nlm.nih.gov/books/NBK395573/

63. Jahangir E, Fazio S, Sampson UKA. Incident diabetes and statins: the blemish of an undisputed heavy weight champion? Br J Clin Pharmacol. 2013 Apr;75(4):955–8.

64. Laakso M, Fernandes Silva L. Statins and risk of type 2 diabetes: mechanism and clinical implications. Front Endocrinol. 2023 Sep 19;14:1239335.

65. Reith C, Preiss D, Blackwell L, Emberson J, Spata E, Davies K, et al. Effects of statin therapy on diagnoses of new-onset diabetes and worsening glycaemia in large-scale randomised blinded statin trials: an individual participant data meta-analysis. Lancet Diabetes Endocrinol. 2024 May 1;12(5):306–19.

66. Goodarzi MO, Li X, Krauss RM, Rotter JI, Chen YDI. Relationship of Sex to Diabetes Risk in Statin Trials. Diabetes Care. 2013 Jul;36(7):e100–1.

67. Culver AL, Ockene IS, Balasubramanian R, Olendzki BC, Sepavich DM, Wactawski-Wende J, et al. Statin Use and Risk of Diabetes Mellitus in Postmenopausal Women in the Women’s Health Initiative. Arch Intern Med. 2012 Jan 23;172(2):144–52.

68. Kao DP, Martin JL, Aquilante CL, Shalowitz EL, Leyba K, Kudron E, et al. Sex-differences in reporting of statin-associated diabetes mellitus to the US Food and Drug Administration. BMJ Open Diabetes Res Care. 2024 Dec 5;12(6):e004343.

69. Benoit S, Ciccia EA, Devarajan P. Cystatin C as a biomarker of chronic kidney disease: latest developments. Expert Rev Mol Diagn. 2020 Oct;20(10):1019–26.

70. West M, Kirby A, Stewart RA, Blankenberg S, Sullivan D, White HD, et al. Circulating Cystatin C Is an Independent Risk Marker for Cardiovascular Outcomes, Development of Renal Impairment, and Long-Term Mortality in Patients With Stable Coronary Heart Disease: The LIPID Study. J Am Heart Assoc. 2022 Mar;11(5):e020745.

71. Maxfield FR, van Meer G. Cholesterol, the central lipid of mammalian cells. Curr Opin Cell Biol. 2010 Aug;22(4):422–9.

72. Werner KB, Elmståhl S, Christensson A, Pihlsgård M. Male sex and vascular risk factors affect cystatin C-derived renal function in older people without diabetes or overt vascular disease. Age Ageing. 2014 May;43(3):411–7.

73. Ambrose JA, Barua RS. The pathophysiology of cigarette smoking and cardiovascular disease: an update. J Am Coll Cardiol. 2004 May 19;43(10):1731–7.

74. Yang Y, Yang C, Lei Z, Rong H, Yu S, Wu H, et al. Cigarette smoking exposure breaks the homeostasis of cholesterol and bile acid metabolism and induces gut microbiota dysbiosis in mice with different diets. Toxicology. 2021 Feb 28;450:152678.

75. Al Musaimi O, Abu-Nawwas AH, Al Shaer D, Khaleel NY, Fawzi M. Influence of age, gender, smoking, diabetes, thyroid and cardiac dysfunctions on cystatin C biomarker. Semergen. 2019;45(1):44–51.

76. Funamoto M, Shimizu K, Sunagawa Y, Katanasaka Y, Miyazaki Y, Komiyama M, et al. Serum Cystatin C, a Sensitive Marker of Renal Function and Cardiovascular Disease, Decreases After Smoking Cessation. Circ Rep. 2019 Nov 30;1(12):623–7.

77. David S, Edwards CW. Forced Expiratory Volume. In: StatPearls [Internet]. Treasure Island (FL): StatPearls Publishing; 2024 [cited 2024 Mar 21]. Available from: http://www.ncbi.nlm.nih.gov/books/NBK540970/

78. Billett HH. Hemoglobin and Hematocrit. In: Walker HK, Hall WD, Hurst JW, editors. Clinical Methods: The History, Physical, and Laboratory Examinations [Internet]. 3rd ed. Boston: Butterworths; 1990 [cited 2025 Apr 4]. Available from: http://www.ncbi.nlm.nih.gov/books/NBK259/

79. Walker HK, Hall WD, Hurst JW, editors. Clinical Methods: The History, Physical, and Laboratory Examinations [Internet]. 3rd ed. Boston: Butterworths; 1990 [cited 2025 Apr 4]. Available from: http://www.ncbi.nlm.nih.gov/books/NBK201/

80. D L. Association between Cigarette Smoking and Serum Gamma-Glutamyl Transferase Level. [cited 2024 Jan 12]; Available from: https://clinmedjournals.org/articles/ijrpm/international-journal-of-respiratory-and-pulmonary-medicine-ijrpm-6-125.php?jid=ijrpm

81. Koenig G, Seneff S. Gamma-Glutamyltransferase: A Predictive Biomarker of Cellular Antioxidant Inadequacy and Disease Risk. Dis Markers [Internet]. 2015 [cited 2024 Jan 12];2015. Available from: https://www.ncbi.nlm.nih.gov/pmc/articles/PMC4620378/

82. Zhang Z, Ma L, Geng H, Bian Y. Effects of Smoking, and Drinking on Serum Gamma-Glutamyl Transferase Levels Using Physical Examination Data: A Cross-Sectional Study in Northwest China. Int J Gen Med. 2021 Apr 15;14:1301–9.

83. Bermudez EA, Rifai N, Buring JE, Manson JE, Ridker PM. Relation between markers of systemic vascular inflammation and smoking in women. Am J Cardiol. 2002 May 1;89(9):1117–9.

84. Singh B, Goyal A, Patel BC. C-Reactive Protein: Clinical Relevance and Interpretation. In: StatPearls [Internet]. Treasure Island (FL): StatPearls Publishing; 2025 [cited 2025 May 8]. Available from: http://www.ncbi.nlm.nih.gov/books/NBK441843/

85. Sonawane MD, Nimse SB. C-Reactive protein: a major inflammatory biomarker. Anal Methods. 2017 Jun 15;9(23):3400–13.

86. Amezcua-Castillo E, González-Pacheco H, Sáenz-San Martín A, Méndez-Ocampo P, Gutierrez-Moctezuma I, Massó F, et al. C-Reactive Protein: The Quintessential Marker of Systemic Inflammation in Coronary Artery Disease—Advancing toward Precision Medicine. Biomedicines. 2023 Sep;11(9):2444.

87. Mehta N, Luthra NS, Corcos DM, Fantuzzi G. C-reactive protein as the biomarker of choice to monitor the effects of exercise on inflammation in Parkinson’s disease. Front Immunol. 2023 May 12;14:1178448.

88. Gallus S, Lugo A, Suatoni P, Taverna F, Bertocchi E, Boffi R, et al. Effect of Tobacco Smoking Cessation on C-Reactive Protein Levels in A Cohort of Low-Dose Computed Tomography Screening Participants. Sci Rep. 2018 Aug 27;8(1):12908.

89. Koh DH, Choi S, Park JH, Lee SG, Kim HC, Kim I, et al. Evaluation on the Sex-Specific Association Between Cigarette Smoke Exposure and Inflammation Markers—C-Reactive Protein and White Blood Cell Count. Nicotine Tob Res. 2024 Apr 1;26(4):484–93.

90. Das I. Raised C-reactive protein levels in serum from smokers. Clin Chim Acta. 1985 Nov 29;153(1):9–13.

91. Yanbaeva DG, Dentener MA, Creutzberg EC, Wesseling G, Wouters EFM. Systemic Effects of Smoking. Chest. 2007 May 1;131(5):1557–66.

92. Tonstad S, Cowan JL. C-reactive protein as a predictor of disease in smokers and former smokers: a review. Int J Clin Pract. 2009 Nov;63(11):1634–41.

93. Espírito Santo LR, Faria TO, Silva CSO, Xavier LA, Reis VC, Mota GA, et al. Socioeconomic status and education level are associated with dyslipidemia in adults not taking lipid-lowering medication: a population-based study. Int Health. 2022 Jul 1;14(4):346–53.

94. Omotoso OE, Teibo JO, Atiba FA, Oladimeji T, Adebesin AO, Babalghith AO. Bridging the genomic data gap in Africa: implications for global disease burdens. Glob Health. 2022 Dec 9;18(1):103.

95. Schoeler T, Speed D, Porcu E, Pirastu N, Pingault JB, Kutalik Z. Participation bias in the UK Biobank distorts genetic associations and downstream analyses. Nat Hum Behav. 2023 Jul;7(7):1216–27.

96. Kelemen LE, Atkinson EJ, de Andrade M, Pankratz VS, Cunningham JM, Wang A, et al. Linkage analysis of obesity phenotypes in pre- and post-menopausal women from a United States mid-western population. BMC Med Genet. 2010 Nov 9;11:156.

97. Ramsey JM, Cooper JD, Penninx BWJH, Bahn S. Variation in serum biomarkers with sex and female hormonal status: implications for clinical tests. Sci Rep. 2016 May 31;6(1):26947.

98. Divani AA, Luo X, Datta YH, Flaherty JD, Panoskaltsis-Mortari A. Effect of Oral and Vaginal Hormonal Contraceptives on Inflammatory Blood Biomarkers. Mediators Inflamm. 2015;2015(1):379501.

99. Schisterman EF, Mumford SL, Sjaarda LA. Failure to Consider the Menstrual Cycle Phase May Cause Misinterpretation of Clinical and Research Findings of Cardiometabolic Biomarkers in Premenopausal Women. Epidemiol Rev. 2014;36(1):71–82.

100. Anckaert E, Jank A, Petzold J, Rohsmann F, Paris R, Renggli M, et al. Extensive monitoring of the natural menstrual cycle using the serum biomarkers estradiol, luteinizing hormone and progesterone. Pract Lab Med. 2021 Mar 13;25:e00211.

101. Zedler BK, Kinser, R., Oey, J., Nelson, B., Roethig, H.-J., Walk, R. A., et al. Biomarkers of exposure and potential harm in adult smokers of 3–7 mg tar yield (Federal Trade Commission) cigarettes and in adult non-smokers. Biomarkers. 2006 Jan 1;11(3):201–20.

